# Mismatches in gene deletions and kidney-related proteins are novel histocompatibility factors in kidney transplantation

**DOI:** 10.1101/2022.02.16.22270977

**Authors:** Salla Markkinen, Ilkka Helanterä, Jouni Lauronen, Marko Lempinen, Jukka Partanen, Kati Hyvärinen

## Abstract

**Background:** The genomic mismatch level between donor and recipient is associated with the risk of acute rejection and long-term graft survival. In the present study, we determined the association of genome-level matching with acute rejection in a single-center kidney transplantation cohort.

**Methods:** Altogether, 1025 pairs of deceased donors and kidney transplant recipients transplanted in 2007–2017 were genotyped. The associations between the sums of whole genome missense variant mismatches and missense mismatches in transmembrane, secretory, and kidney-related proteins, with acute rejection were estimated using Cox proportional hazards model. Additionally, we analyzed 40 common deletion-tagging variants using Cox model.

**Results:** The increasing mismatch sum in kidney-related proteins associated with acute rejection with an unadjusted HR of 1.15 (95% CI, 1.01–1.30; P=0.029) when dividing the sum into quartiles. The sums of other mismatches were not associated with acute rejection. In deletion analysis, a mismatch in rs7542235 (NC_000001.11:g.196854483A>G), genotype GG tagging a homozygous deletion at the complement Factor H-related (CFHR) proteins locus, predisposed to acute rejection with adjusted HR of 2.97 (95% CI, 1.46–6.05; P=0.003).

**Conclusions:** The present study supports that mismatches in gene deletions and kidney-related proteins are novel histocompatibility factors in kidney transplantation. The relative importance of different gene deletions varies between populations, as we found evidence for CFH-related gene deletion but not for the previously reported LIMS1 deletion.

**SIGNIFICANCE STATEMENT:** The first genome-wide association studies have not identified strong candidate genes for complication risks of kidney transplantation whereas matching of non-HLA genes may be promising. The current study determined the association of genome-level matching with acute rejection among 1025 kidney transplant pairs. The increasing mismatch sum in kidney-related proteins was associated with acute allograft rejection. Also, an association between acute allograft rejection and rs7542235 genotype GG, tagging a homozygous deletion at the complement factor H-related (CFHR) loci on chr 1 was found. The present study suggests that mismatches in gene deletions and kidney-related proteins are important novel histocompatibility factors in kidney transplantation.

## INTRODUCTION

Despite a good survival rate of kidney transplants and effective immunosuppressive treatments, up to 10% of recipients worldwide suffer from acute rejection in which the recipient’s immune system recognizes nonself antigens in the allograft and elicits severe alloimmune reaction^1^.

The current histocompatibility matching for transplantation in most centers relies on three major criteria: ABO blood group compatibility, donor-recipient matching at the human leukocyte antigen (HLA) genes, and a cross-matching test to evaluate the preformed antibodies against donor HLA alleles. In genetic terms, transplantation and matching can be regarded as a multifactorial trait in which the matching of HLA and ABO is known to be a critical but clearly not a sufficient factor to fully characterize the risk of acute immunological events.

In recent years, genome-wide association studies (GWAS) for searching novel genetic factors for complications of kidney transplantation have been performed ^2,3,4,5,6,7^. A few genetic associations have been reported ^3,7^ but replication of findings has proven to be difficult. In fact, by far the largest GWAS, including 2094 kidney transplant-pairs with replication in 5866 complete pairs, found no genome-level significant association with graft survival or acute rejection^5^.

An alternative approach to GWAS is to search at the whole genome level for matching of genes or genetic variation that would, in addition to HLA and ABO matching, be important in predicting immunological complications after transplantation. This kind of whole genome-level histocompatibility matching has produced some promising results in hematopoietic stem cell transplantation (HSCT) ^8,9^ and lately in kidney transplantation ^10,11,12,13,14^.

In the present study, we investigated the association between acute rejection and genome-level matching among kidney transplant donor-recipient pairs. We analyzed a retrospective, single-center, deceased donor transplant cohort of 1025 pairs. The results indicate that mismatches in deletions and kidney-related proteins may be novel histocompatibility factors associated with acute rejection.

## METHODS

The study follows the STREGA recommendations (STrengthening the REporting of Genetic Association Studies) in order to enhance the transparency of the report^15^.

### Study population and design

The characteristics of recipients are presented in **Table 1**. The selection of the study cohort is described in **Figure 1**. A total of 1025 kidney transplant recipients older than 18 years who received a first kidney transplantation during 2007–2017 in a single transplant center, at the Helsinki University Hospital, Helsinki, Finland, and 730 HLA-matched adult deceased donors were included in this study. The registration of race and ethnicity data is prohibited in Finland. However, based on principal component analysis, >95% of the study cohort is of Finnish origin. Of the transplanted kidneys, 295 were partner kidneys from one donor to two recipients. The primary outcome examined was biopsy-proven acute rejection, and included both antibody-mediated and T-cell-mediated rejections, defined based on Banff criteria at the time of diagnosis^16^. The borderline rejections were also included in this study. In total, 199 recipients with at least one acute rejection episode and 826 recipients without acute rejection were included.

**Table 1.**
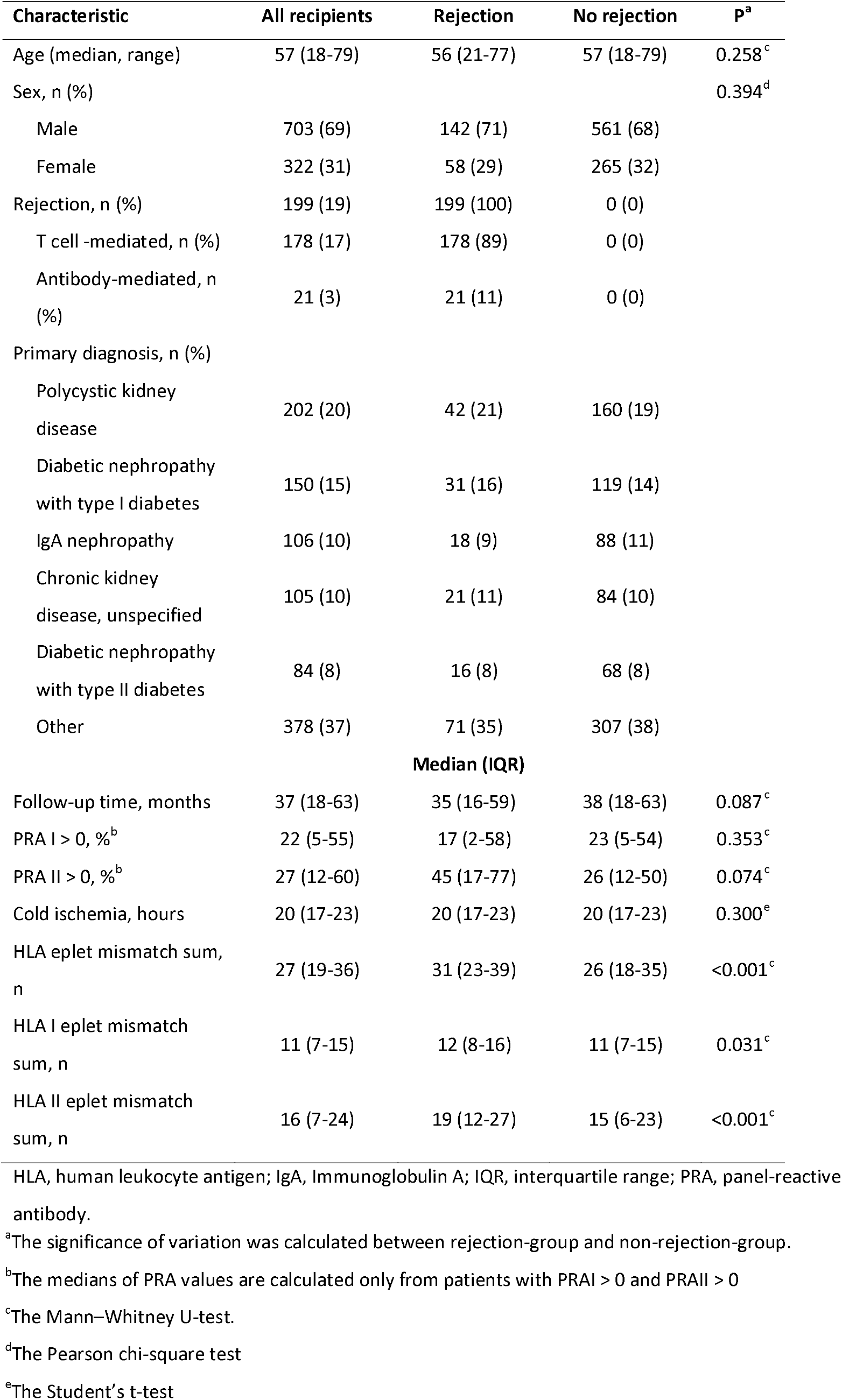
Characteristics of the study population.

**Figure 1.**
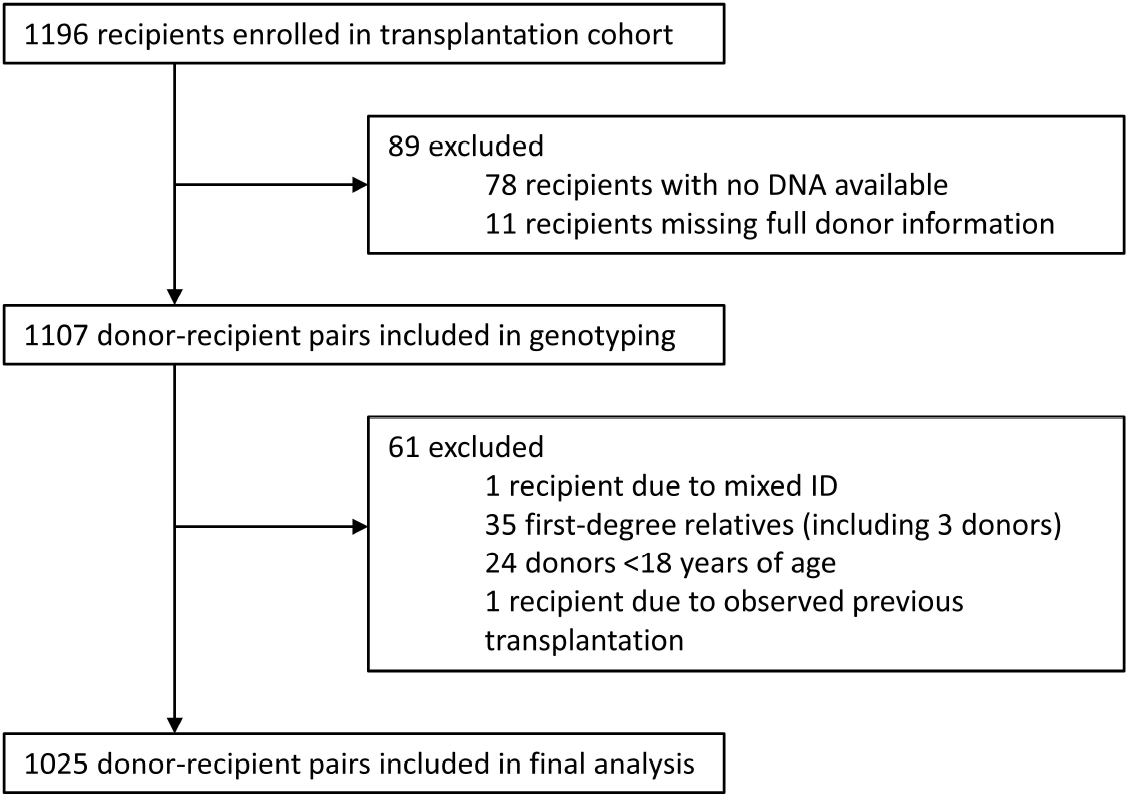
Flow of recipients. The enrolled recipients were ≥18 years of age and received the first kidney from deceased donor between 2007 and 2017.

DNA samples from recipients and donors were extracted from whole blood samples at the time of histocompatibility testing for transplantation at Finnish Red Cross Blood Service, Helsinki, Finland, which is the tissue typing facility for the national transplant center. At organ allocation, the donor-recipient pairs aimed to match the low-resolution level for at the HLA-A, HLA-B and HLA-DRB1 loci, with the DRB1 locus being the most important. All transplantations were negative for complement-dependent cytotoxicity crossmatch test. Panel-reactive antibody (PRA) value for both class I and II was zero in 746/1025 (73%) recipients. PRA I was zero in 788/1025 (77%) and PRA II in 920/1025 (90%) recipients. In the rejection group, 135/199 (68%) recipients were zero for both PRA I and II. PRA I was zero in 145/199 (73%) and PRA II in 172/199 (86%) recipients.

The clinical data of recipients and donors were extracted from the Finnish Transplant Registry, which is a national follow-up registry obliged by law. No loss to follow-up was observed.

The study conforms to the principles of the Declaration of Helsinki and has been approved by the ethics committee of Helsinki University Hospital (HUS/1873/2018) and the Finnish National Supervisory Authority for Welfare and Health (V/9161/2019).

### Genotyping and imputation

Genotyping was performed at the Finnish Institute of Molecular Medicine (FIMM), Helsinki, Finland. The study cohort was genotyped using Illumina’s Infinium Global Screening array-24 v2.0 with multidisease drop-in comprising 759993 variants. The initial quality control removed samples with discordant sex information, duplicate samples, call rate <97%, and heterozygosity excess <-0.3 (not X chromosome) or >0.2.

Prior to imputation, genotyping chip data in GRCh37 build was lifted over to GRCh38/hg38 build following a protocol version 2^17^. The postliftover genotype data were imputed using a Finnish SISu v3 reference panel consisting of high-coverage whole genome sequence (WGS) data from THL Biobank cohorts (N=1768) (Pärn *et al*. manuscript in preparation). The imputation procedure followed the genotype imputation pipeline version 2^18^. The flow of genetic variants after imputation is presented in **Figure 2**. After imputation, 32321074 variants were available for quality control procedures. Individuals with a missing genotype >5%, variants with minor allele frequency (MAF) <1% and variants with a missing data rate >10% were excluded from the postimputation quality control (PLINK 1.9^19^). Additionally, variants with an INFO score of <0.6 were excluded. After postimputation filtering, 9085413 variants were available for the analyses. PLINK IBD analysis was performed to identify relatives, and 35 first-degree relatives were excluded from the study cohort.

**Figure 2.**
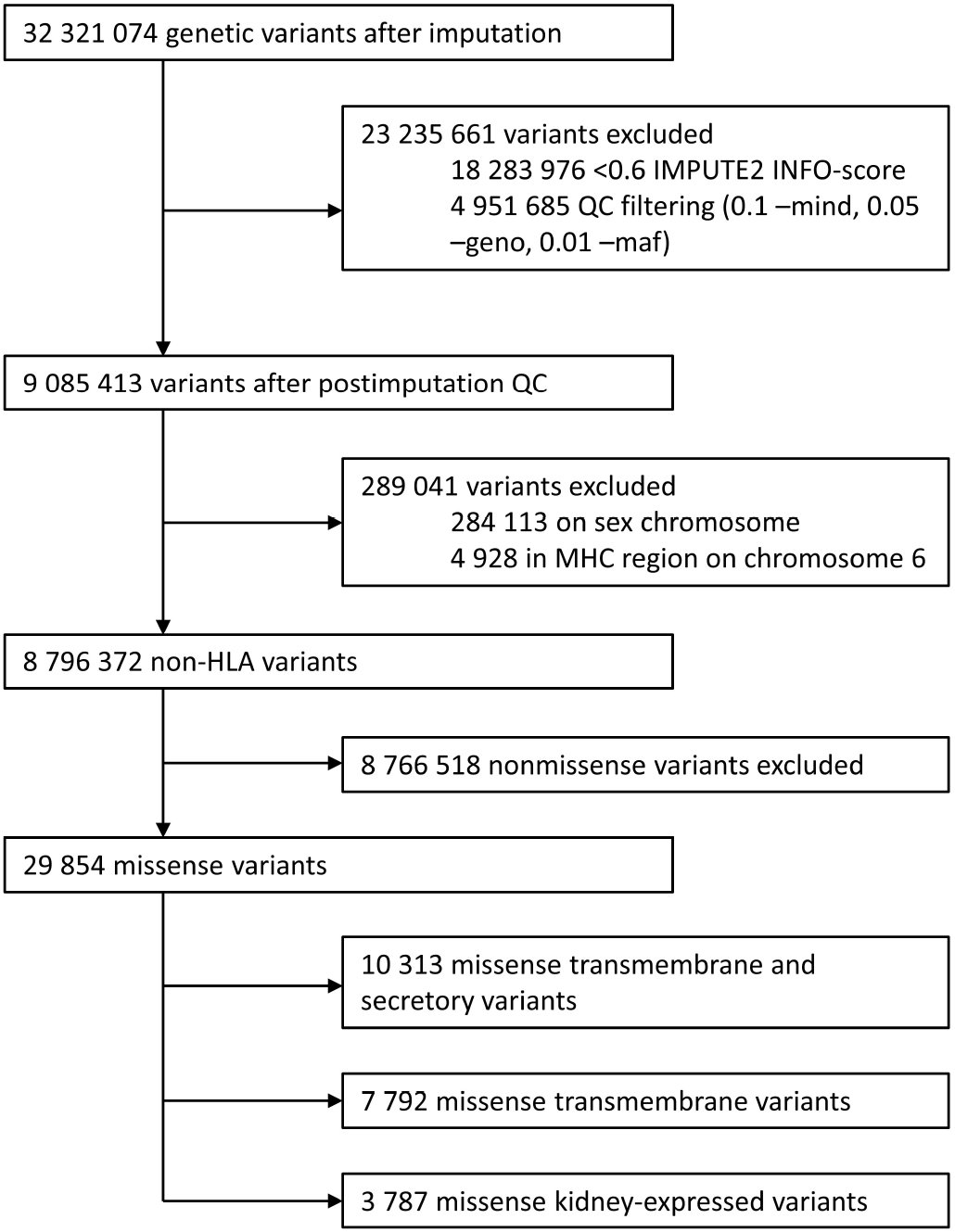
Flow of genetic variants.

In addition to the clinical low-resolution HLA typing of the individual samples, HLA type was imputed to high resolution using HIBAG v1.0.3 with the Finnish HLA reference for HLA-A, HLA-B, HLA-C, HLA-DRB1, HLA-DPB1, HLA-DQA1 and DQB1 gene alleles^20^.

### Genome-wide mismatch analyses

We carried out a replication analysis of a recently reported mismatch study by Reindl-Schwaighofer *et al*.^12^. The HLA eplet mismatch was calculated based on imputed high resolution HLA genotypes using HLAMatchmaker^21^.

The online tool Ensemble Variant Effect Predictor release 103 (https://www.ensembl.org/Homo_sapiens/Tools/VEP) was used for functional annotation of observed and imputed variants^22^. **Figure 2** summarizes the flow of genetic variants. To retrieve only missense variants, we used the filtering operator “consequence is missense_variant”. Transcripts for transmembrane and secreted proteins, transmembrane proteins and kidney-related proteins were retrieved from UniProt (https://www.uniprot.org/)^23^. The query string (annotation:(type:transmem) OR locations:(location:”Secreted [SL-0243]”) OR keyword:”Transmembrane [KW-0812]” AND reviewed:yes) AND organism:”Homo sapiens (Human) [9606]” was used to identify transmembrane and secretory proteins. The query string (annotation:(type:transmem) OR keyword:”Transmembrane [KW-0812]” AND reviewed:yes) AND organism:”Homo sapiens (Human) [9606]” was used to identify transmembrane proteins. Additionally to the Reindl-Schwaighofer *et al*.^12^ protocol we used query string (annotation:(type:”tissue specificity” kidney) AND reviewed:yes) AND organism:”Homo sapiens (Human) [9606]” to retrieve the information for kidney-related proteins. Ensembl transcript IDs linked to proteins returned by these query strings were retrieved. The transcript IDs from UniProt and VEP were merged to create the final missense variant lists of proteins of interest.

The variant mismatch was defined as the donor carrying an allele that was not present in the recipient. We calculated the sum of missense variant mismatches across the genome between donor and recipient for transmembrane and secreted proteins, transmembrane proteins and kidney-related proteins using R v3.6.2. Additionally, we calculated the overall missense mismatch sum (i.e., not limiting to only transmembrane and secretory proteins). Because of the adjustment for HLA eplet mismatch, the variants in the major histocompatibility complex (MHC) region (28,510,120–33,480,577) on chromosome 6 and on sex chromosomes were excluded from the present study.

We evaluated the association of missense variant mismatch with acute rejection using logistic regression with recipient and donor sex, recipient and donor age, cold ischemia time, PRA I, PRA II, HLA I (HLA-A, -B, C) and HLA II (HLA-DRB1, -DQA1, -DQB1 and -DPB1) eplet mismatch as covariates. The missense mismatch sum was also tested in a univariate model that was not adjusted for additional covariates. In addition, we carried out a multivariable Cox proportional hazards model against time to acute rejection with recipient and donor sex, recipient and donor age, cold ischemia time, PRA I, PRA II, and HLA I and HLA II eplet mismatch as covariates. Two individuals were excluded because of missing follow-up data. Recipients were also divided into quartiles based on their mismatch sum, and the relationship of each quartile with time to acute rejection was evaluated with Kaplan–Meier curves and tested with both Cox proportional hazards and logistic regression models.

### Deletion analysis

A recent study by Steers *et al*.^13^ reported a significant association with allograft rejection in the LIMS1 gene deletion. We carried out a replication study, and a total of 40 deletion-tagging variants were available in our dataset (Table S1 in the Supplementary data). The tagging variants analyzed had a global MAF of more than 10%, and were all in strong linkage disequilibrium with the deletions^13^. We tested the deletion-tagging variants in our cohort of 1025 donor-recipient pairs, and additionally, evaluated the variants in recipient-only data.

In the deletion analyses, the dependent variable was time-to-first-rejection, defined from the date of first transplant to the date of rejection event. We used Kaplan–Meier survival curves and Cox proportional hazards models to calculate the estimates for both the recipient-donor pairs and recipients only. In the recipient-donor analysis (collision model), the risk group was defined when a recipient who was homozygous for a deletion-tagging variant received a kidney from a donor who was either nonhomozygous or homozygous for the reference variant. The mismatch status was used as an independent variable. The mismatch statuses for all deletions are reported in Table S1. In the Cox proportional hazards model the other covariates were recipient and donor sex, recipient and donor age, cold ischemia time and HLA I and HLA II eplet mismatch. Two individuals were excluded because of missing follow-up data. We conducted a Bonferroni-correction threshold for statistical significance (level of 0.05/40, or 1.25−10^−3^).

We calculated the mismatch sum of all homozygous deletions in donor-recipient pairs among the 40 deletion-tagging variants and evaluated the association of mismatch sum to time to acute rejection with Kaplan–Meier and Cox proportional hazards models. The sum of all homozygous deletions in recipient-only data was also calculated, and the association was assessed with time to acute rejection. The associations of deletion sum with acute rejection outcomes in both models were also evaluated with logistic regression.

### Whole genome sequencing

To validate the deletions in complement factor H (CFH) region, whole genome sequencing (WGS) was performed on three recipients homozygous for CFH region deletion-tagging variant rs7542235. Sequencing was performed at FIMM, Helsinki, Finland, and the samples were sequenced with Illumina’s NovaSeq S4 NS4-300 run.

After Illumina’s NovaSeq S4 NS4-300 run, the total number of aligned bases for each of the three samples was ∼110 gb. The input reads for each sample were approximately 1 billion, and the length for each read was 151 base pairs. The coverage depth for our samples was 34–36x. General statistics for the WGS run are presented in Supplementary Table S2.

### Detection of CFH and CFH antibodies

For determination of CFH and to assess de novo antibody formation against CFH, blood serum samples were used in enzyme-linked immunosorbent assays (ELISAs). We analyzed all serum samples available for the CFHR deletion homozygous patients and some control samples (**Table 2**).

**Table 2.** The number of pre- and post-transplantation serum samples in CFH- and CFH-IgG ELISA.

| Deletion genotype | Rejection status | CFH ELISA |  | CFH IgG ELISA |  |
| --- | --- | --- | --- | --- | --- |
|  |  | Pre-tx serum sample (n) | Post-tx serum sample (n) | Pre-tx serum sample (n) | Post-tx serum sample (n) |
| Deletion-homozygous | Rejection | 5 | 5 | 5 | 5 |
| Deletion-homozygous | No rejection | 5 | 1 | 5 | 1 |
| Non-risk genotype | Rejection | 10 | 10 | 10 | 10 |
| Non-risk genotype | No rejection | 10 | 6 | 30 | 26 |
| aHUS, non-risk genotype | No rejection | 2 | 1 | 2 | 1 |
| In total |  | 32 | 23 | 52 | 43 |

aHUS, atypical hemolytic uremic syndrome; CFH, complement factor H; ELISA, enzyme-linked immunosorbent assay; IgG, Immunoglobulin G; tx, transplantation.

In total, 55 samples were analyzed for CFH expression (**Table 2**). Detection of CFH from serum samples was determined by using CFH ELISA kits (Abnova, Taipei, Taiwan) according to the manufacturer’s instructions. A dilution 1:200000 was used for each sample.

The presence of antibodies against CFH was determined by using CFH IgG ELISA kits (Abnova, Taipei, Taiwan) according to the manufacturer’s recommendations, with a 1:50 dilution of samples. In the CFH IgG ELISA, in total of 95 serum samples were analyzed (**Table 2**).

### Other statistical analyses

Characteristics of recipients were nonnormally distributed and thus described by medians and ranges, medians and interquartile ranges (IQRs) for continuous variables, and frequencies and percentages for binary variables (**Table 1**). The significance of variation between characteristics in the two cohorts (rejection and nonrejection) was analyzed using the nonparametric Mann-Whitney U-test for nonnormally distributed data (recipient age, follow-up time, PRA I and II, HLA eplet mismatch, HLA I eplet mismatch and HLA II eplet mismatch), Pearson’s chi-square test for categorical data (recipient sex), or Student’s t-test for normally distributed data (cold ischemia). P values <0.05 were considered statistically significant.

We assessed the association of clinical covariates with acute rejection using a logistic regression model (Supplementary Table S3). P values <0.05 were considered statistically significant. No missing data was observed.

Statistical tests were carried out in R v3.6.2 using the R function glm for logistic regression and survival packages for survival analyses.

As we had only a few relevant cases in ELISA analyses to compare the differences between pre- and posttransplantation serum samples, we did not perform any statistical tests.

## RESULTS

### Analysis of characteristics and covariates

The clinical characteristics are compared between the rejection and nonrejection groups in **Table 1**. A total of 199 rejection events were available in the present cohort of 1025 recipients. Of these, 178 were T-cell-mediated and 21 antibody-mediated rejections. The median follow-up time of the cohort was 37 months (IQR 16–59 months). The overall HLA eplet mismatch and HLA II eplet mismatch sums were significantly different between the groups with a P value <0.001.

Donor sex, donor age, recipient age, PRA II and mismatches in HLA II between donor and recipient emerged from the logistic regression model as significant predictors of acute rejection (Supplementary Table S4). Female donor sex was associated with an increased risk of acute rejection, as well as older donor age, younger recipient age and an increase in PRA II value and HLA II mismatch sum.

### Whole-genome mismatch analyses

After postimputation filtering and exclusion of MHC regions, sex chromosomes and nonmissense variants, 29854 missense variants were available for analysis at the whole genome level. A total of 10313 of the variants were located in transmembrane or secretory proteins, 7792 in transmembrane proteins and 3787 in kidney-related proteins (**Figure 2**).

The median number of mismatches between donor and recipient in transmembrane and secretory proteins was 1765 (IQR 1724–1812). For the transmembrane proteins the median mismatch sum was 1334 (IQR 1292–1671), and for kidney-related proteins, it was 605 (IQR 585–627). The overall genome-wide missense variant mismatch sum was 4935 (IQR 4861– 5012).

No evidence for a whole-genome-level association of missense variants with acute rejection was found in either the Cox proportional hazards or logistic regression model (Supplementary Tables S4-S5). We replicated a missense mismatch study of transmembrane and secretory proteins as reported by Reindl-Schwaighofer *et al*.^12^ and a missense mismatch study of kidney-related proteins. Our data show that increasing mismatch sum in kidney-related proteins increases the risk for acute rejection with an unadjusted HR of 1.15 (95% CI, 1.01–1.30, P=0.029) when dividing the mismatch sum into quartiles (**Figure 3A**).

**Figure 3.**
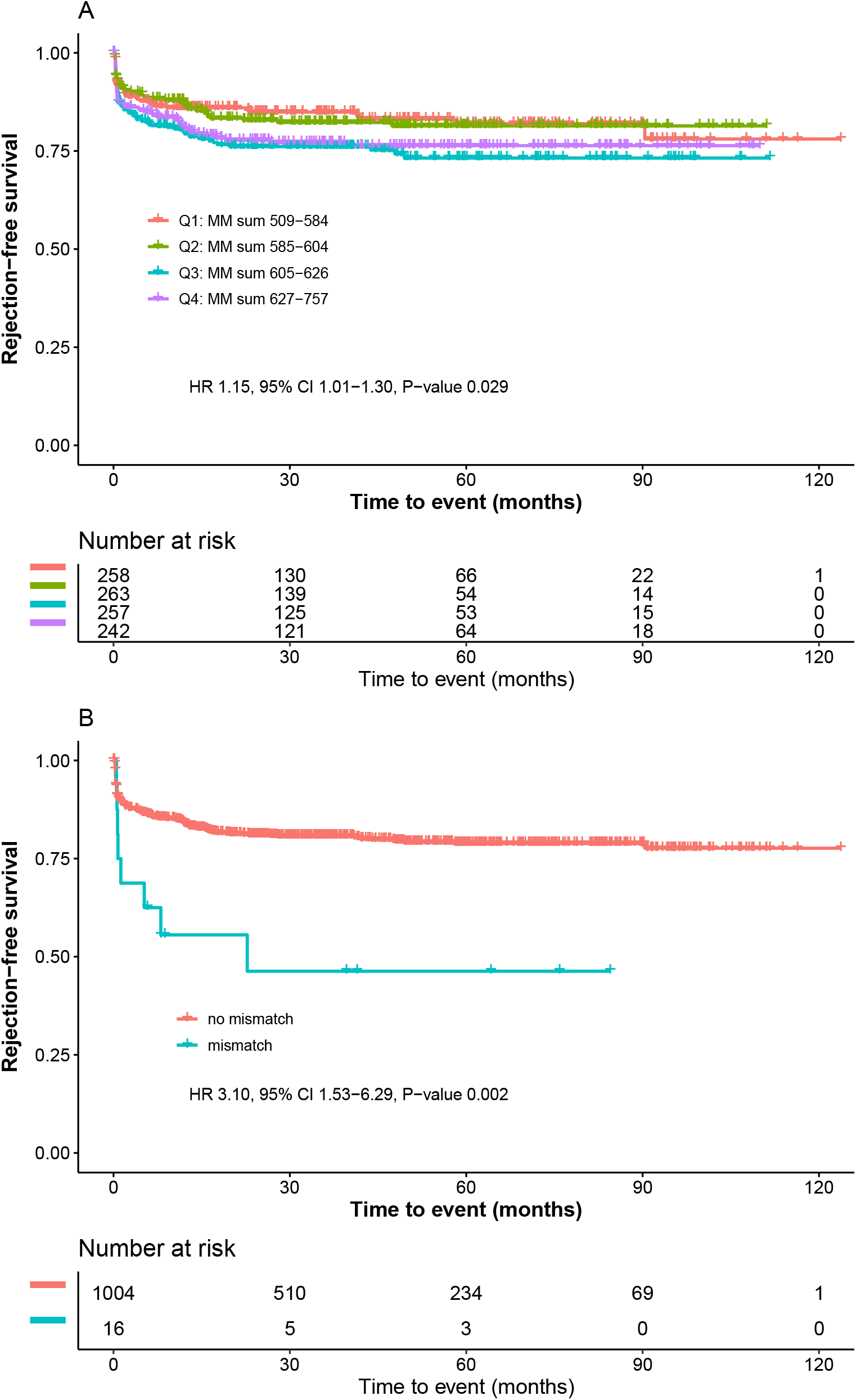
The effect of quartiles of missense variant mismatch sum coding for kidney-related proteins and rs7542235 mismatch on rejection-free graft survival in recipient-donor pairs. A) The effect of quartiles of missense variant mismatch sum coding for kidney-related proteins on rejection-free graft survival. The quartile 1 (Q1) represents the lowest number of mismatches between recipient and donor, quartile 4 (Q4) the highest number of mismatches. Unadjusted hazard ratio (HR) with confidence interval (CI) shown. B) The effect of deletion-tagging variant rs7542235 mismatch on rejection-free graft survival in recipient-donor pairs. In recipient-donor analysis the recipient who was homozygous for a deletion-tagging allele G received a transplant from a donor with AG or AA genotype. The orange curve represents the no mismatch status and the light blue curve represents the mismatch status. An event (acute rejection) occurs each time the curve drops. The tick marks indicate censored data (end of follow-up time). Unadjusted hazard ratio (HR) with confidence interval (CI) shown. CI, confidence interval; HR, hazard ratio; MM, mismatch

We did not find a statistically significant association between the missense mismatch sum of transmembrane and secretory proteins and time to acute rejection in the multivariable Cox proportional hazards model (Supplementary Table S6) or in the logistic regression model (Supplementary Table S7). The other missense mismatch sums of transmembrane or kidney-related proteins were not significant predictors of time to acute rejection or acute rejection outcome in our data (Supplementary Tables S8-S11).

In the Kaplan–Meier and Cox proportional hazards model analyses of the quartiles of mismatch sum, we found no significance in the subsets of transmembrane and secretory proteins, transmembrane proteins or in all missense variants and acute rejection (Supplementary Figure S1-S3).

### Deletion analysis

In total, 40 of the deletion-tagging variants analyzed by Steers *et al*. were available in the present dataset (Table S1 in the Supplementary data), and they were tested in the donor-recipient mismatch analysis^13^. A mismatch refers to cases in which a recipient who is homozygous for a deletion-tagging variant received a transplant from a donor with a nonhomozygous or homozygous reference variant genotype. We also tested the variants in recipient-only analysis.

The rs7542235 genotype GG has been reported to tag for deletions in CFH region, namely CFH-related proteins (CFHR) 1-3^24,25^. We observed that mismatch in rs7542235 was significantly associated with a higher risk for rejection than the no mismatch status with unadjusted HR of 3.10 (95% CI, 1.53–6.29; P=0.002) and adjusted HR of 2.97 (95% CI, 1.46– 6.05; P=0.003) (**Table 3**). The number of recipients with rs7542235 genotype GG in rejection group was 8 (4%) and in non-rejection group it was 8 (1%), and none of their donors were homozygous for the deletion-tagging variant. **Figure 3B** shows the Kaplan–Meier plot of the effect of the deletion-tagging variant rs7542235 on rejection-free graft survival. The statistical significance of the association, however, fails to pass the Bonferroni-corrected threshold of 0.00125. The same results were also observed in recipient-only data. No other deletion-tagging variants showed an association with these outcomes.

**Table 3.**
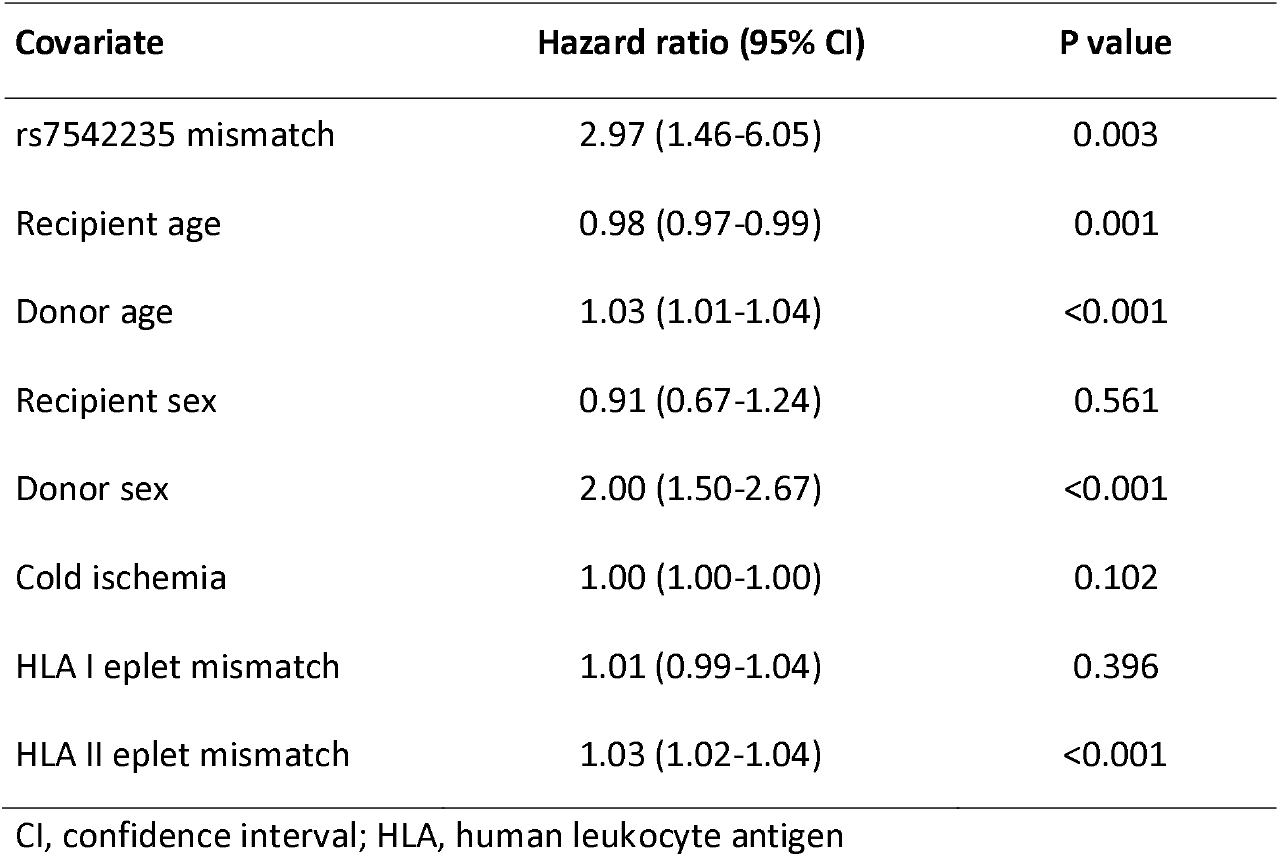
Cox proportional hazards model on the association of rs7542235 mismatch to time to acute rejection.

| **Covariate** | **Hazard ratio (95% CI)** | **P value** |
| --- | --- | --- |
| rs7542235 mismatch | 2.97 (1.46-6.05) | 0.003 |
| Recipient age | 0.98 (0.97-0.99) | 0.001 |
| Donor age | 1.03 (1.01-1.04) | <0.001 |
| Recipient sex | 0.91 (0.67-1.24) | 0.561 |
| Donor sex | 2.00 (1.50-2.67) | <0.001 |
| Cold ischemia | 1.00 (1.00-1.00) | 0.102 |
| HLA I eplet mismatch | 1.01 (0.99-1.04) | 0.396 |
| HLA II eplet mismatch | 1.03 (1.02-1.04) | <0.001 |

CI, confidence interval; HLA, human leukocyte antigen

The study by Steers *et al*. reported a significant association with allograft rejection when recipients had LIMS1 gene deletion^13^. We carried out a replication study in both recipient-donor and recipient-only settings but could not find evidence for the association of LIMS1 deletion or other deletion-tagging variants when using the Kaplan–Meier curve and Cox proportional hazards model (Supplementary Table S12-S13).

The median number of all homozygous deletions in donor-recipient pairs was 3 (IQR 2-4, range 0-9), and in recipient-only data, it was 4 (IQR 3-5, range 0-10). We did not find a significant association between the deletion sum and acute rejection in between donor-recipient pairs or among recipients (Supplementary Figure S4-S5, Supplementary Table S14-S17).

### Whole genome sequencing

Whole-genome sequencing confirmed that deletion-tagging variant rs7542235 is located at the position of chr1:196,854,483 between CFHR1 and CFHR4. The genotype GG tags a homozygous deletion at the CFHR1 locus on chr 1. Each of the three GG recipients sequenced had a homozygous deletion of different sizes at the CFHR1 region, and two of them also had a homozygous deletion completely or partly at the CFHR3 loci (Supplementary Figure S6).

### Detection of CFH and CFH antibodies

The results for the ELISA analyses are presented in Supplementary data (Supplementary Table S18 and S19). All serum samples from 55 recipients, including 16 with homozygous deletion at the CFHR1 locus, showed normal levels of CFH in ELISA analysis. Both pre- and postserum samples were available from only 5 recipients with homozygous deletion who received a transplant from a nondeletion donor. We were not able to find evidence for anti-CFH antibody formation between pre- and postserum samples. Only one deletion-homozygous recipient with acute rejection showed an increase in anti-CFH antibody in postserum sample.

## DISCUSSION

The results of the current study indicate that mismatches in gene deletions and kidney-related proteins may predispose to acute rejection and present novel histocompatibility factors in kidney transplantation. The analysis was conducted in a large single-center and single-population cohort of 1025 kidney transplantation recipient-donor pairs, reducing the impact of confounding factors. In the study, we investigated to what extent differences or matching between donor-recipient pairs (i) at the overall genome level, (ii) in graft-expressed proteins, and (iii) in common gene deletions were associated with the increased risk for acute rejection. The current gold standard for matching recipients and donors in transplantations relies on HLA and ABO matching and negative HLA crossmatching. Modern genome tools enable us to expand the analysis of all genetic variation between transplant pairs and address the role of non-HLA variation. In principle, we can assume that any immunogenic protein-level difference found in donors and missing in recipients can lead to alloimmune reactions. There is indeed emerging evidence that genetic differences between recipient and donor, in e.g., graft- or cell surface-expressed proteins or even at the overall genomic level, correlate with the risk of transplantation complications such as acute rejection or long-term graft loss^10,11,12,13,14^.

Reindl-Schwaighofer *et al*. reported a statistically significant association in unadjusted Cox proportional hazards model between a transmembrane and secretory nonsynonymous donor-recipient mismatch sum and time to ten years and death censored graft failure in a cohort of 477 transplant pairs^12^. We did not find a similar association in the present cohort. However, our results showed that increasing mismatch sum in kidney-related proteins was associated with acute rejection when dividing the sum into quartiles in survival analysis. This endpoint is not identical to that of Reindl-Schwaighofer but may reflect a similar long-term effect since the acute rejection has also been associated with a decrease in long-term survival. The study of Mesnard *et al*. also found an association between long-term outcome and overall genetic mismatch score. It is plausible that the overall alloantigenic load of the donor graft is associated with long-term rather than acute effects that may be more readily handled by medication^10^.

Steers *et al*. investigated the association of common deletion-tagging variants and kidney allograft rejection^13^. The researchers hypothesized, along with the concept introduced by McCarroll *et al*. in the HSCT setting, that a recipient whose genome lacks a kidney-related gene product due to a gene deletion should raise alloimmune reaction to graft from a donor whose genome carries the functional gene, hence expressing the protein^8^. Steers *et al*. found that kidney recipients with two copies of a deletion near a gene called LIMS1 had a significantly higher risk of rejection when the donor had at least one functional copy of the same gene. In the present study cohort, we were not able to replicate the finding of the LIMS1 deletion. We instead found an association between an acute rejection and another deletion-tagging variant rs7542235. This variant tags a deletion at the CFHR1-3 region on chromosome 1. rs7542235 GG-homozygotes are assumed to be homozygotes for deletion of these genes. We confirmed the tagging by genomic sequencing: three rs7542235 GG-homozygote recipients had a homozygous deletion in the CFHR locus. Interestingly, the deletions were not identical in size and encompassed different genes. The homozygous deletion in the CFHR locus was statistically significantly associated with a decreasing probability of rejection-free graft survival in recipient-donor pairs. The same finding was seen in recipient-only data as well, most likely as it is highly unlikely that an unrelated organ donor would have the same deletion. In fact, if deletions indeed prove to be novel risk markers for transplantation complications as now indicated by a few studies including the present study^8,13^, it may be possible to estimate the complication risk due to gene deletions directly from patient genome data and before transplantation if unrelated donors are used.

To further study the effect of CFHR1-locus deletion on the protein level and its possible alloantigenic role, we performed anti-CFH antibody ELISA for pre- and posttransplantation serum samples of 5 deletion homozygotes. In this small set of samples, we could not find evidence for anti-CFH formation post transplantation in deletion homozygotes. It is not clear whether the antibody response is raised against other proteins expressed by the genes located in the CFHR1 locus. Previous studies have shown that anti-CFH antibody formation is strongly associated with CFHR1 and CFHR3 deletions^26,27,28^, and later is has been suggested that the deletion of CFHR1 alone could be involved in anti-CFH antibody formation^27^. Additionally, the CFH levels did not differ between the nonrisk genotype and deletion genotype. This result was expected since the CFH gene was intact based on the whole sequencing results.

Complement Factor H is a control protein of the complement system, inhibiting the alternative pathway of complement activation. Depending on the individual CFHR genes, it has been associated with a number of diseases, including atypical hemolytic uremic syndrome (aHUS), C3 glomerulopathies, IgA nephropathy, age related macular degeneration and systemic lupus erythematosus^26,27,28,29,30^. The functions of CFHR1 and CFHR3 are still unclear, but these two proteins have been suggested to be complement regulators as well^29^. It has been proposed that complement activity is determined by a homeostatic balance between CFHR1, CFHR3 and CFH^30^. The present finding of the deletion may be related to both primary kidney disease and outcome of transplantation.

There are some limitations in the present study. The significance of the kidney-related mismatch analysis was lost after adjusting the data with additional covariates (Supplementary Table S20). The p value of our findings in the deletion mismatch study did not achieve the Bonferroni corrected level of significance. The Bonferroni correction, however, can be very conservative when several tests are included. In the deletion mismatch analysis, the frequency of risk variant, rs7542235 genotype GG, was very low in the study population with total cases of 16 out of 1025 (2%) individuals. Additionally, the ideal control group for a deletion mismatch would be those patients who were deletion homozygous and received a deletion-matched graft. Unfortunately, such cases were very low in numbers or not found at all.

We had access to only a few pre- and posttransplantation samples and furthermore were not able to estimate the quality of serum samples because they were primarily collected for crossmatch testing and stored in the freezer for even longer periods of time. Clearly, a prospective collection of serum samples from deletion homozygotes should be performed. In the present study, we were able to replicate two previous recipient-donor mismatch studies, but clearly larger, prospective collaborative studies should be performed to obtain sufficient power for genome-level studies in organ transplantation.

In summary, we found an association between the increasing mismatch sum in kidney-related proteins and acute rejection. Sums of overall genomic missense mismatches or mismatches in transmembrane and secretory proteins were not associated with acute rejection. Additionally, we conclude that common gene deletions may play an important role in novel histocompatibility in kidney transplantation. As we were not able to repeat the previous finding of LIMS1 gene deletion, the relative importance of different gene deletions appears to vary between populations.

## Data Availability

The code for variant selection and statistical testing is publicly available in GitHub (https://github.com/FRCBS/Kidney_genome_wide_mismatch). Genetic data are not publicly available due to restrictions issued by the ethical committee and current legislation in Finland that do not allow the distribution of pseudonymized personal data, including genetic and clinical data.

## ABBREVIATIONS

aHUS: atypical hemolytic uremic syndrome
CFH: complement Factor H
CFHR: complement Factor H-related protein
CFHR1: complement Factor H-related protein 1
CFHR3: complement Factor H-related protein 3
CFHR4: complement Factor H-related protein 4
CI: confidence interval
ELISA: enzyme-linked immunosorbent array
FIMM: Finnish Institute for Molecular Medicine
GWAS: genome-wide association study
HLA: human leukocyte antigen
HR: hazard ratio
HSCT: hematopoietic stem cell transplantation
IgA: Immunoglobulin A
IgG: Immunoglobulin G
IQR: interquartile range
MHC: major histocompatibility complex
MM: mismatch
OR: odds ratio
PRA: panel-reactive antibody
SNP: single nucleotide polymorphism
TX: transplantation
VEP: variant effect predictor
WGS: whole genome sequencing

## AUTHOR CONTRIBUTION

SM, JP, KH planned the study, interpreted the results and prepared the manuscript; SM and KH carried out the genome data analysis; IH, ML, JL collected the clinical data; all authors read, accepted and contributed to the final version of the manuscript.

## ACKNOWLEDGEMENTS

We want to thank Drs Satu Koskela, Jarmo Ritari and Mikko Arvas for their help, Ms Sisko Lehmonen for help in collecting the samples and the staff of the HLA laboratory for collaboration. The data used for the research was imputed with the THL Biobank’s SISu v3 Imputation reference panel obtained from THL Biobank. We thank all study participants for their generous participation in the FINRISK, Health 2000 and Migraine Family studies. We also thank the Sequencing Informatics Team, FIMM Human Genomics, University of Helsinki for the work done in preparation of the reference panel data.

## SUPPORTING INFORMATION STATEMENT

Additional supporting information may be found online in the Supporting Information section.

