## Supplementary data for "Mismatches in gene deletions and kidney-related proteins are novel histocompatibility factors in kidney transplantation"

**Table of contents**

Supplemental Tables	4

Supplementary Table S1. Deletion-tagging variants and their frequencies in the study cohort	4

Supplementary Table S2. General statistics of the whole genome sequencing with Illumina’s NovaSeq S4 NS4-300 run	6

Supplementary Table S3. Logistic regression model on the association of clinical covariates and acute rejection	7

Supplementary Table S4. Cox proportional hazards model on the association of mismatch sum of all missense variants to time to acute rejection	8

Supplementary Table S5. Logistic regression model on the association of mismatch sum of all missense variants and acute rejection	9

Supplementary Table S6. Cox proportional hazards model on the association of mismatch sum of secretory and transmembrane variants to time to acute rejection	10

Supplementary Table S7. Logistic regression model on the association of mismatch sum of secretory and transmembrane variants and acute rejection	11

Supplementary Table S8. Cox proportional hazards model on the association of mismatch sum of transmembrane variants to time to acute rejection	12

Supplementary Table S9. Logistic regression model on the association of mismatch sum of transmembrane variants and acute rejection	13

Supplementary Table S10. Cox proportional hazards model on the association of mismatch sum of kidney-related variants to time to acute rejection	14

Supplementary Table S11. Logistic regression model on the association of mismatch sum of kidney-related variants and acute rejection	15

Supplementary Table S12. Cox proportional hazards model on the association of all deletion-tagging variant mismatches to time to acute rejection	16

Supplementary Table S13. Cox proportional hazards model on the association of all deletion-tagging variants to time to acute rejection in recipient data.	17

Supplementary Table S14. Cox proportional hazards model on the association of the sum of deletion mismatches between donor-recipient to time to acute rejection	18

Supplementary Table S15. Logistic regression model for the sum of deletion mismatches between donor-recipient pairs and acute rejection	19

Supplementary Table S16. Cox proportional hazards model on the association of the sum of deletions and time to acute rejection in recipient data	20

Supplementary Table S17. Logistic regression model for the sum of deletions and acute rejection in recipient data	21

Supplementary Table S18. CFH ELISA results for serum samples	22

Supplementary Table S19. CFH IgG ELISA results for serum samples	24

Supplementary Table S20. Cox proportional hazards model on the association of quartiles of mismatch sum of kidney-related variants and acute rejection	27

Supplemental Figures	28

Supplementary Figure S1. The effect of quartiles of missense variant mismatch sum coding for transmembrane and secretory proteins on rejection-free graft survival	28

Supplementary Figure S2. The effect of quartiles of missense variant mismatch sum coding for transmembrane proteins on rejection-free graft survival	29

Supplementary Figure S3. The effect of quartiles of the overall genome-wide missense variant mismatch sum on rejection-free graft survival	30

Supplementary Figure S4. The effect of deletion mismatch sum on rejection-free graft survival in donor-recipient pairs	31

Supplementary Figure S5. The effect of deletion sum on rejection-free graft survival among recipients	32

Supplementary Figure S6. The results of whole genome sequencing of CFH- and CFH-related protein loci on chromosome 1	33

**Supplementary Table S1. Deletion-tagging variants and their frequencies** **in the study cohort.**

| **CHR** | **Deletion-tagging variant** | **DNA Variant HGVS Nomenclature** | **A1** | **A2** | **MAF** | **Mismatches^a^ (%)** | | **Affected genes^b^** |
| --- | --- | --- | --- | --- | --- | --- | --- | --- |
|  |  |  |  |  |  | **No rejection group**  **(n = 826)** | **Rejection group**  **(n = 199)** |  |
| 1 | rs10927864 | NC_000001.11:g.15839166C>G | G | C | 0.27 | 58 (7) | 14 (7) | RPS16P1 |
| 1 | rs11209948 | NC_000001.11:g.72346221G>T | G | T | 0.36 | 83 (10) | 23 (12) | RPL31P12 |
| 1 | rs11249248 | NC_000001.11:g.25426560T>C | T | C | 0.46 | 135 (16) | 38 (19) | LOC100288240/RHCE/SDHDP7/TMEM50A |
| 1 | rs11587012 | NC_000001.11:g.152786728A>C | C | A | 0.28 | 65 (8) | 12 (6) | LCE1D/LCE1E/LOC100289267 |
| 1 | rs158736 | NC_000001.11:g.222193326G>C | C | G | 0.34 | 88 (11) | 18 (9) | LOC100287974 |
| 1 | rs6693105 | NC_000001.11:g.152618187T>C | T | C | 0.37 | 94 (11) | 24 (12) | LCE3B/LCE3C |
| 1 | rs7542235 | NC_000001.11:g.196854483A>G | G | A | 0.14 | 8 (1) | 8 (4) | CFH/CFHR1/CFHR3/LOC100289145 |
| 2 | rs7419565 | NC_000002.12:g.130195449T>C | C | T | 0.41 | 110 (13) | 19 (10) | TUBA3E |
| 2 | rs893403 | NC_000002.12:g.108606280G>A | G | A | 0.31 | 66 (8) | 13 (7) | LOC100288532 |
| 5 | rs10053292 | NC_000005.10:g.149896561T>C | C | T | 0.11 | 4 (0.5) | 2 (1) | HMGXB3/PDE6A/RPS20P4/SLC26A2/TIGD6 |
| 5 | rs2387715 | NC_000005.10:g.180934266A>T | T | A | 0.31 | 92 (11) | 21 (11) | BTNL3/BTNL8/LOC100128762/LOC646227 |
| 5 | rs7703761 | NC_000005.10:g.148177325C>T | C | T | 0.45 | 133 (16) | 35 (18) | SPINK5L2 |
| 6 | rs17654108 | NC_000006.12:g.121480387A>T | T | A | 0.12 | 15 (2) | 3 (2) | LOC260339 |
| 7 | rs2160195 | NC_000007.14:g.38365370A>T | T | A | 0.34 | 92 (11) | 22 (11) | TRGV1/TRGV2/TRGV3/TRGV4/TRGV5 |
| 7 | rs4621754 | NC_000007.14:g.158706303A>G | G | A | 0.09 | 7 (1) | 2 (1) | NCAPG2 |
| 7 | rs4729606 | NC_000007.14:g.100724167T>C | C | T | 0.26 | 44 (5) | 16 (8) | ZAN |
| 7 | rs6943474 | NC_000007.14:g.101357735A>G | G | A | 0.47 | 165 (20) | 34 (17) | EMID2 |
| 8 | rs11985201 | NC_000008.11:g.39581402G>A | A | G | 0.42 | 115 (14) | 33 (17) | ADAM3A/ADAM5P |
| 8 | rs4543566 | NC_000008.11:g.6968014C>G | G | C | 0.12 | 15 (2) | 4 (2) | DEFA10P |
| 9 | rs1523688 | NC_000009.12:g.104592374T>G | G | T | 0.24 | 52 (6) | 8 (4) | OR13C2/OR13C5 |
| 9 | rs2174926 | NC_000009.12:g.118763272A>G | A | G | 0.42 | 136 (16) | 26 (13) | LOC442434 |
| 10 | rs10885336 | NC_000010.11:g.112351444G>A | A | G | 0.38 | 113 (14) | 22 (11) | GUCY2G |
| 10 | rs2342606 | NC_000010.11:g.80034407T>C | T | C | 0.44 | 140 (17) | 27 (14) | LOC642521/LOC642538 |
| 10 | rs3793917 | NC_000010.11:g.122459759C>G | G | C | 0.26 | 49 (6) | 20 (10) | ARMS2 |
| 11 | rs11228868 | NC_000011.10:g.55252994C>T | T | C | 0.07 | 2 (0.2) | 1 (0.5) | TRIM48 |
| 11 | rs1944862 | NC_000011.10:g.55521460G>A | A | G | 0.31 | 70 (8) | 21 (11) | OR4P1P |
| 11 | rs4882017 | NC_000011.10:g.48548630A>G | A | G | 0.34 | 215 (26) | 52 (26) | OR4A45P |
| 12 | rs1478309 | NC_000012.12:g.10389146T>G | T | G | 0.19 | 32 (4) | 7 (4) | KLRC1/KLRC2/KLRC3 |
| 13 | rs9318648 | NC_000013.11:g.24566498A>G | A | G | 0.29 | 60 (7) | 18 (9) | LOC374491 |
| 14 | rs11156875 | NC_000014.9:g.35150610A>G | G | A | 0.19 | 29 (4) | 5 (3) | RPL23AP70 |
| 14 | rs8007442 | NC_000014.9:g.21924807T>C | T | C | 0.44 | 127 (15) | 28 (14) | TRAV14DV4 |
| 14 | rs8022070 | NC_000014.9:g.81411038C>T | T | C | 0.12 | 14 (2) | 6 (3) | LOC731308 |
| 16 | rs10521145 | NC_000016.10:g.28585563G>A | A | G | 0.09 | 8 (1) | 1 (0.5) | SULT1A1 |
| 16 | rs2244613 | NC_000016.10:g.55810697G>T | G | T | 0.19 | 31 (4) | 8 (4) | CES4 |
| 17 | rs16966699 | NC_000017.11:g.41343914C>G | G | C | 0.11 | 11 (1) | 2 (1) | KRT33A, KRT33B |
| 17 | rs8064493 | NC_000017.11:g.41237071A>G | A | G | 0.20 | 27 (3) | 9 (5) | KRTAP9P1 |
| 19 | rs103294 | NC_000019.10:g.54293995T>C | T | C | 0.29 | 61 (7) | 23 (12) | LILRA3 |
| 19 | rs324121 | NC_000019.10:g.52391192G>A | A | G | 0.12 | 13 (2) | 1 (0.5) | LOC400713 |
| 19 | rs3810336 | NC_000019.10:g.56175525G>A | A | G | 0.28 | 64 (8) | 14 (7) | GALP |
| 19 | rs4806152 | NC_000019.10:g.35395758A>C | C | A | 0.22 | 38 (5) | 5 (3) | FFAR3/GPR42P |

A1, minor allele; A2, major allele; CHR, chromosome; MAF, minor allele frequency

**^a^**Mismatch refers to cases in which a recipient who is homozygous for a deletion-tagging allele received a transplant from a donor with a nonhomozygous or homozygous reference allele genotype.

**^b^**Steers NJ, Li Y, Drace Z, et al. Genomic Mismatch at LIMS1 Locus and Kidney Allograft Rejection. *N Engl J Med*. 2019;380(20):1918-1928. doi:10.1056/nejmoa1803731

**Supplementary Table S2. General statistics of the whole genome sequencing with Illumina’s NovaSeq S4 NS4-300 run.**

| **Sample Name** | **Total number of variants** | **Sex** | **Total number of aligned reads** | **Total number of aligned bases** | **Coverage depth** | **Percentage of sites in region with at least 20x coverage** | **Total number of input reads, millions** | **% of unmapped reads** | **% of properly paired reads** | **Median insert size** |
| --- | --- | --- | --- | --- | --- | --- | --- | --- | --- | --- |
| JUIHLJXRHX | 4935191 | XX | 723.8 | 110081.9 | 36.1 x | 93.3 % | 1039.2 | 0.9% | 97.5% | 393 |
| QDM2EBUWCF | 4957367 | XX | 698.7 | 106263.2 | 34.9 x | 93.2 % | 976.9 | 1.1% | 97.3% | 413 |
| V53C3LDXJW | 4882956 | XY | 736.6 | 112034.0 | 36.7 x | 91.7 % | 1045.4 | 1.2% | 97.2% | 405 |

**Supplementary Table S3. Logistic regression model on the association of clinical covariates and acute rejection.**

| **Covariate** | **Odds ratio (95% CI)** | **P value** |
| --- | --- | --- |
| Recipient sex | 0.78 (0.52-1.13) | 0.192 |
| Donor sex | 2.23 (1.60-3.11) | <0.001 |
| Recipient age | 0.98 (0.96-0.99) | 0.002 |
| Donor age | 1.03 (1.01-1.05) | 0.001 |
| Cold ischemia | 1.00 (1.00-1.00) | 0.395 |
| PRAI | 1.00 (0.99-1.01) | 0.957 |
| PRAII | 1.01 (1.00-1.02) | 0.011 |
| HLAI eplet mismatch | 1.01 (0.98-1.04) | 0.436 |
| HLAII eplet mismatch | 1.03 (1.02-1.05) | <0.001 |

CI, confidence interval; HLA, human leukocyte antigen; PRA, panel reactive antibody

**Supplementary Table S4. Cox proportional hazards** **model on the association of mismatch sum of all missense variants to time to acute rejection.**

| **Covariate** | **Hazard ratio (95% CI)** | **P value** |
| --- | --- | --- |
| Mismatch sum of all missense variants | 1.00 (1.00-1.00) | 0.465 |
| Recipient age | 0.98 (0.97-0.99) | 0.001 |
| Donor age | 1.03 (1.01-1.04) | <0.001 |
| Recipient sex | 0.80 (0.57-1.12) | 0.194 |
| Donor sex | 1.99 (1.49-2.66) | <0.001 |
| Cold ischemia | 1.00 (1.00-1.00) | 0.118 |
| PRAI | 1.00 (0.99-1.01) | 0.544 |
| PRAII | 1.01 (1.00-1.02) | 0.007 |
| HLAI eplet mismatch | 1.01 (0.99-1.03) | 0.453 |
| HLAII eplet mismatch | 1.03 (1.02-1.04) | <0.001 |

CI, confidence interval; HLA, human leukocyte antigen; PRA, panel reactive antibody

**Supplementary Table S5. Logistic regression model on the association of mismatch sum of all missense variants and acute rejection.**

| **Covariate** | **Odds ratio (95% CI)** | **P value** |
| --- | --- | --- |
| Mismatch sum of all missense variants | 1.00 (1.00-1.00) | 0.632 |
| Recipient sex | 0.78 (0.53-1.13) | 0.192 |
| Donor sex | 2.23 (1.61-3.11) | <0.001 |
| Recipient age | 0.98 (0.96-0.99) | 0.001 |
| Donor age | 1.03 (1.01-1.05) | 0.001 |
| Cold ischemia | 1.00 (1.00-1.00) | 0.396 |
| PRAI | 1.00 (0.99-1.01) | 0.980 |
| PRAII | 1.01 (1.00-1.02) | 0.011 |
| HLAI eplet mismatch | 1.01 (0.98-1.04) | 0.423 |
| HLAII eplet mismatch | 1.04 (1.02-1.05) | <0.001 |

CI, confidence interval; HLA, human leukocyte antigen; PRA, panel reactive antibody

**Supplementary Table S6. Cox proportional hazards model on the association of mismatch sum of secretory and transmembrane variants to time to acute rejection.**

| **Covariate** | **Hazard ratio (95% CI)** | **P value** |
| --- | --- | --- |
| Mismatch sum of secretory and transmembrane variants | 1.00 (1.00-1.00) | 0.581 |
| Recipient age | 0.98 (0.97-0.99) | 0.001 |
| Donor age | 1.03 (1.01-1.04) | <0.001 |
| Recipient sex | 0.80 (0.57-1.12) | 0.193 |
| Donor sex | 1.98 (1.48-2.65) | <0.001 |
| Cold ischemia | 1.00 (1.00-1.00) | 0.118 |
| PRAI | 1.00 (0.99-1.01) | 0.546 |
| PRAII | 1.01 (1.00-1.02) | 0.007 |
| HLAI eplet mismatch | 1.01 (0.99-1.03) | 0.451 |
| HLAII eplet mismatch | 1.03 (1.02-1.04) | <0.001 |

CI, confidence interval; HLA, human leukocyte antigen; PRA, panel reactive antibody

**Supplementary Table S7. Logistic regression model on the association of mismatch sum of secretory and transmembrane variants and acute rejection.**

| **Covariate** | **Odds ratio (95% CI)** | **P value** |
| --- | --- | --- |
| Mismatch sum of secretory and transmembrane variants | 1.00 (1.00-1.00) | 0.673 |
| Recipient sex | 0.78 (0.53-1.13) | 0.192 |
| Donor sex | 2.23 (1.61-3.10) | <0.001 |
| Recipient age | 0.98 (0.96-0.99) | 0.001 |
| Donor age | 1.03 (1.01-1.05) | 0.001 |
| Cold ischemia | 1.00 (0.99-1.00) | 0.402 |
| PRAI | 1.00 (0.99-1.01) | 0.978 |
| PRAII | 1.01 (1.00-1.02) | 0.011 |
| HLAI eplet mismatch | 1.01 (0.98-1.04) | 0.420 |
| HLAII eplet mismatch | 1.04 (1.02-1.05) | <0.001 |

CI, confidence interval; HLA, human leukocyte antigen; PRA, panel reactive antibody

**Supplementary Table S8. Cox proportional hazards model on the association of mismatch sum of transmembrane variants to time to acute rejection.**

| **Covariate** | **Hazard ratio (95% CI)** | **P value** |
| --- | --- | --- |
| Mismatch sum of transmembrane variants | 1.00 (1.00-1.00) | 0.393 |
| Recipient age | 0.98 (0.97-0.99) | 0.001 |
| Donor age | 1.03 (1.01-1.04) | <0.001 |
| Recipient sex | 0.80 (0.57-1.12) | 0.189 |
| Donor sex | 1.98 (1.48-2.64) | <0.001 |
| Cold ischemia | 1.00 (1.00-1.00) | 0.119 |
| PRAI | 1.00 (0.99-1.01) | 0.541 |
| PRAII | 1.01 (1.00-1.02) | 0.007 |
| HLAI eplet mismatch | 1.01 (0.99-1.03) | 0.435 |
| HLAII eplet mismatch | 1.03 (1.02-1.04) | <0.001 |

CI, confidence interval; HLA, human leukocyte antigen; PRA, panel reactive antibody

**Supplementary Table S9. Logistic regression model on the association of mismatch sum of transmembrane variants and acute rejection.**

| **Covariate** | **Odds ratio (95% CI)** | **P value** |
| --- | --- | --- |
| Mismatch sum of transmembrane variants | 1.00 (1.00-1.00) | 0.485 |
| Recipient sex | 0.77 (0.52-1.13) | 0.189 |
| Donor sex | 2.22 (1.60-3.10) | <0.001 |
| Recipient age | 0.98 (0.96-0.99) | 0.001 |
| Donor age | 1.03 (1.01-1.05) | 0.001 |
| Cold ischemia | 1.00 (1.00-1.00) | 0.403 |
| PRAI | 1.00 (0.99-1.01) | 0.985 |
| PRAII | 1.01 (1.00-1.02) | 0.010 |
| HLAI eplet mismatch | 1.01 (0.98-1.04) | 0.410 |
| HLAII eplet mismatch | 1.04 (1.02-1.05) | <0.001 |

CI, confidence interval; HLA, human leukocyte antigen; PRA, panel reactive antibody

**Supplementary Table S10. Cox proportional hazards model on the association of mismatch sum of kidney-related variants to time to acute rejection.**

| **Covariate** | **Hazard ratio (95% CI)** | **P value** |
| --- | --- | --- |
| Mismatch sum of kidney-related variants | 1.00 (1.00-1.01) | 0.409 |
| Recipient age | 0.98 (0.97-0.99) | 0.001 |
| Donor age | 1.03 (1.01-1.04) | <0.001 |
| Recipient sex | 0.80 (0.58-1.13) | 0.204 |
| Donor sex | 1.98 (1.48-2.65) | <0.001 |
| Cold ischemia | 1.00 (1.00-1.00) | 0.104 |
| PRAI | 1.00 (0.99-1.01) | 0.582 |
| PRAII | 1.01 (0.98-1.03) | 0.010 |
| HLAI eplet mismatch | 1.01 (0.98-1.03) | 0.483 |
| HLAII eplet mismatch | 1.03 (1.02-1.04) | <0.001 |

CI, confidence interval; HLA, human leukocyte antigen; PRA, panel reactive antibody

**Supplementary Table S11. Logistic regression model on the association of mismatch sum of kidney-related variants and acute rejection.**

| **Covariate** | **Odds ratio (95% CI)** | **P value** |
| --- | --- | --- |
| Mismatch sum of kidney-related variants | 1.00 (1.00-1.01) | 0.258 |
| Recipient sex | 0.78 (0.53-1.14) | 0.204 |
| Donor sex | 2.22 (1.60-3.10) | <0.001 |
| Recipient age | 0.98 (0.96-0.99) | 0.001 |
| Donor age | 1.03 (1.01-1.05) | 0.001 |
| Cold ischemia | 1.00 (1.00-1.00) | 0.364 |
| PRAI | 1.00 (0.99-1.01) | 0.924 |
| PRAII | 1.01 (1.00-1.02) | 0.012 |
| HLAI eplet mismatch | 1.01 (0.98-1.04) | 0.432 |
| HLAII eplet mismatch | 1.03 (1.02-1.05) | <0.001 |

CI, confidence interval; HLA, human leukocyte antigen; PRA, panel reactive antibody

**Supplementary Table S12. Cox proportional hazards model on the association of all deletion-tagging variant mismatches to time to acute rejection.**

|  |  | **Unadjusted** |  | **Adjusted** |  |
| --- | --- | --- | --- | --- | --- |
| **CHR** | **Tagging variant** | **Hazard Ratio**  **(95 % CI)** | **P-Value** | **Hazard Ratio**  **(95 % CI)** | **P-Value** |
| 1 | rs10927864 | 1.09 (0.63–1.87) | 0.76 | 1.12 (0.65–1.93) | 0.69 |
| 1 | rs11209948 | 1.16 (0.75–1.78) | 0.52 | 1.26 (0.81–1.94) | 0.31 |
| 1 | rs11249248 | 1.21 (0.85–1.73) | 0.28 | 1.36 (0.95–1.95) | 0.09 |
| 1 | rs11587012 | 0.78 (0.43–1.39) | 0.39 | 0.74 (0.41–1.33) | 0.32 |
| 1 | rs158736 | 0.88 (0.55–1.44) | 0.62 | 0.97 (0.59–1.57) | 0.89 |
| 1 | rs6693105 | 1.10 (0.72–1.69) | 0.65 | 1.10 (0.71–1.70) | 0.66 |
| 1 | rs7542235 | 3.10 (1.53–6.29) | 0.002 | 2.97 (1.46–6.05) | 0.003 |
| 2 | rs7419565 | 0.71 (0.44–1.13) | 0.15 | 0.71 (0.44–1.14) | 0.15 |
| 2 | rs893403 | 0.88 (0.50–1.54) | 0.64 | 0.85 (0.48–1.50) | 0.58 |
| 5 | rs10053292 | 1.54 (0.38–6.21) | 0.54 | 2.01 (0.50–8.13) | 0.33 |
| 5 | rs2387715 | 0.96 (0.61–1.50) | 0.85 | 1.08 (0.69–1.71) | 0.73 |
| 5 | rs7703761 | 1.15 (0.80–1.65) | 0.46 | 1.12 (0.78–1.62) | 0.54 |
| 6 | rs17654108 | 0.87 (0.28–2.72) | 0.81 | 0.90 (0.29–2.83) | 0.86 |
| 7 | rs2160195 | 1.01 (0.65–1.58) | 0.10 | 0.89 (0.57–1.39) | 0.61 |
| 7 | rs4621754 | 1.29 (0.32–5.19) | 0.72 | 1.07 (0.26–4.36) | 0.92 |
| 7 | rs4729606 | 1.37 (0.82–2.29) | 0.23 | 1.19 (0.71–2.00) | 0.50 |
| 7 | rs6943474 | 0.88 (0.61–1.27) | 0.48 | 0.89 (0.61–1.29) | 0.53 |
| 8 | rs11985201 | 1.20 (0.83–1.74) | 0.34 | 1.18 (0.81–1.71) | 0.39 |
| 8 | rs4543566 | 1.05 (0.39–2.82) | 0.93 | 0.96 (0.35–2.60) | 0.94 |
| 9 | rs1523688 | 0.69 (0.34–1.40) | 0.30 | 0.77 (0.38–1.56) | 0.47 |
| 9 | rs2174926 | 0.78 (0.51–1.17) | 0.23 | 0.85 (0.56–1.28) | 0.43 |
| 10 | rs10885336 | 0.79 (0.51–1.24) | 0.31 | 0.76 (0.49–1.19) | 0.23 |
| 10 | rs2342606 | 0.81 (0.54–1.22) | 0.32 | 0.90 (0.60–1.35) | 0.60 |
| 10 | rs3793917 | 1.60 (1.01–2.55) | 0.05 | 1.40 (0.88–2.25) | 0.16 |
| 11 | rs11228868 | 1.52 (0.21–10.8) | 0.68 | 1.31 (0.18–9.47) | 0.80 |
| 11 | rs1944862 | 1.24 (0.79–1.95) | 0.35 | 1.20 (0.76–1.89) | 0.43 |
| 11 | rs4882017 | 0.98 (0.72–1.35) | 0.10 | 0.96 (0.70–1.32) | 0.79 |
| 12 | rs1478309 | 0.93 (0.44–1.98) | 0.90 | 0.77 (0.36–1.64) | 0.50 |
| 13 | rs9318648 | 1.22 (0.75–1.98) | 0.42 | 1.18 (0.73–1.92) | 0.51 |
| 14 | rs11156875 | 0.70 (0.29–1.71) | 0.44 | 0.76 (0.31–1.86) | 0.55 |
| 14 | rs8007442 | 0.92 (0.62–1.37) | 0.69 | 0.98 (0.65–1.46) | 0.90 |
| 14 | rs8022070 | 1.64 (0.73–3.69) | 0.23 | 1.46 (0.65–3.31) | 0.36 |
| 16 | rs10521145 | 0.48 (0.07–3.43) | 0.47 | 0.49 (0.07–3.53) | 0.48 |
| 16 | rs2244613 | 1.13 (0.56–2.29) | 0.74 | 0.84 (0.41–1.72) | 0.63 |
| 17 | rs16966699 | 0.79 (0.20–3.17) | 0.74 | 1.24 (0.30–5.03) | 0.78 |
| 17 | rs8064493 | 1.33 (0.68–2.60) | 0.40 | 1.33 (0.68–2.61) | 0.41 |
| 19 | rs103294 | 1.59 (1.03–2.46) | 0.04 | 1.53 (0.99–2.38) | 0.06 |
| 19 | rs324121 | 0.32 (0.04–2.25) | 0.25 | 0.41 (0.06–2.94) | 0.38 |
| 19 | rs3810336 | 0.93 (0.54–1.60) | 0.79 | 0.94 (0.54–1.64) | 0.83 |
| 19 | rs4806152 | 0.59 (0.24–1.44) | 0.25 | 0.66 (0.27–1.60) | 0.36 |

CHR, chromosome

**Supplementary Table S13. Cox proportional hazards model on the association of all deletion-tagging variants to time to acute rejection in recipient data.**

|  |  | **Unadjusted** |  | **Adjusted** |  |
| --- | --- | --- | --- | --- | --- |
| **CHR** | **Tagging variant** | **Hazard Ratio**  **(95 % CI)** | **P-Value** | **Hazard Ratio**  **(95 % CI)** | **P-Value** |
| 1 | rs10927864 | 1.21 (0.73–2.02) | 0.47 | 1.24 (0.74–2.08) | 0.41 |
| 1 | rs11209948 | 1.49 (1.03–2.16) | 0.03 | 1.50 (1.04–2.17) | 0.03 |
| 1 | rs11249248 | 1.22 (0.88–1.69) | 0.23 | 1.35 (0.97–1.87) | 0.07 |
| 1 | rs11587012 | 0.85 (0.49–1.47) | 0.56 | 0.82 (0.48–1.42) | 0.49 |
| 1 | rs158736 | 0.95 (0.60–1.49) | 0.83 | 1.01 (0.64–1.60) | 0.95 |
| 1 | rs6693105 | 0.95 (0.62–1.46) | 0.82 | 0.95 (0.60–1.44) | 0.75 |
| 1 | rs7542235 | 3.10 (1.53–6.29) | 0.002 | 2.97 (1.46–6.05) | 0.003 |
| 2 | rs7419565 | 0.73 (0.47–1.11) | 0.14 | 0.69 (0.45–1.06) | 0.09 |
| 2 | rs893403 | 0.81 (0.46–1.43) | 0.47 | 0.78 (0.45–1.38) | 0.40 |
| 5 | rs10053292 | 1.54 (0.38–6.21) | 0.54 | 2.01 (0.50–8.13) | 0.33 |
| 5 | rs2387715 | 0.95 (0.61–1.48) | 0.83 | 1.08 (0.69–1.69) | 0.73 |
| 5 | rs7703761 | 1.01 (0.72–1.42) | 0.96 | 1.00 (0.71–1.41) | 0.10 |
| 6 | rs17654108 | 0.87 (0.28–2.72) | 0.81 | 0.90 (0.29–2.83) | 0.86 |
| 7 | rs2160195 | 1.25 (0.85–1.85) | 0.26 | 1.14 (0.77–1.69) | 0.52 |
| 7 | rs4621754 | 1.29 (0.32–5.19) | 0.72 | 1.07 (0.26–4.36) | 0.92 |
| 7 | rs4729606 | 1.41 (0.87–2.29) | 0.17 | 1.27 (0.78–2.07) | 0.34 |
| 7 | rs6943474 | 0.93 (0.67–1.27) | 0.64 | 0.93 (0.67–1.28) | 0.65 |
| 8 | rs11985201 | 1.10 (0.77–1.57) | 0.60 | 1.09 (0.76–1.56) | 0.64 |
| 8 | rs4543566 | 1.05 (0.39–2.82) | 0.93 | 0.96 (0.35–2.60) | 0.94 |
| 9 | rs1523688 | 0.63 (0.31–1.28) | 0.20 | 0.70 (0.35–1.43) | 0.33 |
| 9 | rs2174926 | 0.75 (0.51–1.10) | 0.14 | 0.79 (0.53–1.17) | 0.24 |
| 10 | rs10885336 | 0.88 (0.59–1.32) | 0.54 | 0.84 (0.56–1.27) | 0.41 |
| 10 | rs2342606 | 0.88 (0.61–1.27) | 0.49 | 0.95 (0.66–1.38) | 0.80 |
| 10 | rs3793917 | 1.56 (0.98–2.48) | 0.06 | 1.39 (0.87–2.22) | 0.17 |
| 11 | rs11228868 | 1.52 (0.21–10.8) | 0.68 | 1.31 (0.18–9.47) | 0.79 |
| 11 | rs1944862 | 1.20 (0.77–1.87) | 0.42 | 1.16 (0.74–1.81) | 0.52 |
| 11 | rs4882017 | 0.97 (0.73–1.28) | 0.81 | 0.97 (0.73–1.28) | 0.81 |
| 12 | rs1478309 | 1.16 (0.60–2.27) | 0.66 | 0.95 (0.48–1.86) | 0.88 |
| 13 | rs9318648 | 1.24 (0.78–1.97) | 0.36 | 1.17 (0.74–1.86) | 0.50 |
| 14 | rs11156875 | 0.64 (0.26–1.55) | 0.32 | 0.70 (0.29–1.70) | 0.43 |
| 14 | rs8007442 | 0.92 (0.64–1.32) | 0.65 | 0.97 (0.67–1.40) | 0.86 |
| 14 | rs8022070 | 1.90 (0.89–4.03) | 0.10 | 1.70 (0.79–3.63) | 0.17 |
| 16 | rs10521145 | 0.48 (0.07-3.43) | 0.47 | 0.49 (0.07–3.53) | 0.48 |
| 16 | rs2244613 | 1.09 (0.54–2.21) | 0.81 | 0.82 (0.40–1.68) | 0.59 |
| 17 | rs16966699 | 0.79 (0.20–3.17) | 0.74 | 1.24 (0.30–5.03) | 0.77 |
| 17 | rs8064493 | 1.33 (0.68–2.60) | 0.40 | 1.33 (0.68–2.61) | 0.41 |
| 19 | rs103294 | 1.53 (0.99–2.36) | 0.06 | 1.49 (0.69–2.30) | 0.08 |
| 19 | rs324121 | 0.29 (0.04–2.09) | 0.22 | 0.39 (0.05–2.80) | 0.35 |
| 19 | rs3810336 | 0.90 (0.53–1.53) | 0.70 | 0.91 (0.53–1.55) | 0.73 |
| 19 | rs4806152 | 0.53 (0.22–1.29) | 0.16 | 0.61 (0.25–1.48) | 0.27 |

CHR, chromosome

**Supplementary Table S14. Cox proportional hazards model on the association of the sum of deletion mismatches between donor-recipient to time to acute rejection.**

| **Covariate** | **Hazard ratio (95% CI)** | **P value** |
| --- | --- | --- |
| Deletion sum mismatches | 1.02 (0.94-1.10) | 0.699 |
| Recipient age | 0.98 (0.97-0.99) | 0.002 |
| Donor age | 1.03 (1.01-1.04) | <0.001 |
| Recipient sex | 0.92 (0.67-1.25) | 0.576 |
| Donor sex | 2.01 (1.50-2.69) | <0.001 |
| Cold ischemia | 1.00 (1.00-1.00) | 0.111 |
| HLAI eplet mismatch | 1.01 (0.98-1.03) | 0.466 |
| HLAII eplet mismatch | 1.03 (1.02-1.04) | <0.001 |

CI, confidence interval; HLA, human leukocyte antigen

**Supplementary Table S15. Logistic regression model for the sum of deletion mismatches between donor-recipient pairs and acute rejection.**

| **Covariate** | **Odds ratio (95% CI)** | **P value** |
| --- | --- | --- |
| Deletion sum mismatches | 1.01 (0.92-1.12) | 0.761 |
| Recipient sex | 0.87 (0.61-1.24) | 0.445 |
| Donor sex | 2.25 (1.63-3.14) | <0.001 |
| Recipient age | 0.98 (0.96-0.99) | 0.002 |
| Donor age | 1.03 (1.01-1.05) | 0.001 |
| Cold ischemia | 1.00 (1.00-1.00) | 0.357 |
| HLAI eplet mismatch | 1.01 (0.98-1.04) | 0.437 |
| HLAII eplet mismatch | 1.04 (1.02-1.05) | <0.001 |

CI, confidence interval; HLA, human leukocyte antigen

**Supplementary Table S16. Cox proportional hazards model on the association of the sum of deletions and time to acute rejection in recipient data.**

| **Covariate** | **Hazard ratio (95% CI)** | **P value** |
| --- | --- | --- |
| Deletion sum | 1.02 (0.95-1.10) | 0.571 |
| Recipient age | 0.98 (0.97-0.99) | 0.002 |
| Donor age | 1.03 (1.01-1.04) | <0.001 |
| Recipient sex | 0.92 (0.67-1.25) | 0.578 |
| Donor sex | 2.01 (1.50-2.69) | <0.001 |
| Cold ischemia | 1.00 (1.00-1.00) | 0.111 |
| HLAI eplet mismatch | 1.01 (0.98-1.03) | 0.471 |
| HLAII eplet mismatch | 1.03 (1.02-1.04) | <0.001 |

CI, confidence interval; HLA, human leukocyte antigen

**Supplementary Table S17. Logistic regression model for the sum of deletions and acute rejection in recipient data.**

| **Covariate** | **Odds ratio (95% CI)** | **P value** |
| --- | --- | --- |
| Deletion sum | 1.02 (0.93-1.11) | 0.677 |
| Recipient sex | 0.87 (0.61-1.24) | 0.444 |
| Donor sex | 2.25 (1.62-3.13) | <0.001 |
| Recipient age | 0.98 (0.96-0.99) | 0.002 |
| Donor age | 1.03 (1.01-1.05) | 0.001 |
| Cold ischemia | 1.00 (1.00-1.00) | 0.360 |
| HLAI eplet mismatch | 1.01 (0.98-1.04) | 0.438 |
| HLAII eplet mismatch | 1.04 (1.02-1.05) | <0.001 |

CI, confidence interval; HLA, human leukocyte antigen

**Supplementary Table S18. CFH ELISA results for serum samples**

| **Sample ID** | **Pre/post** | **CFH (µg/mL)** |
| --- | --- | --- |
| ABGR4XFUNH | pre | 380,4 |
| ABGR4XFUNH | post | 792,4 |
| BCZORKR4A3 | pre | 453,4 |
| DGYMLR5QCF | pre | 813,2 |
| EN257NXZIH | pre | 612,2 |
| NQZTUVJ6JA | pre | 648,6 |
| 3RVWTIW47P | pre | 260,9 |
| 3RVWTIW47P | post | 954,8 |
| ADCKGCMCUK | pre | 820,3 |
| ADCKGCMCUK | post | 421,9 |
| JUIHLJXRHX | pre | 733,0 |
| JUIHLJXRHX | post | 570,7 |
| QDM2EBUWCF | pre | 347,5 |
| QDM2EBUWCF | post | 542,8 |
| V53C3LDXJW | pre | 544,9 |
| V53C3LDXJW | post | 306,0 |
| TSMOSHTLU4 | pre | 411,9 |
| TSMOSHTLU4 | post | 630,0 |
| ADSAZ3KKXD | pre | 437,6 |
| 2C26AIVPU6 | pre | 524,2 |
| 2C26AIVPU6 | post | 721,6 |
| 2LQUST2UOF | pre | 836,8 |
| 32DVSNKGWW | pre | 948,4 |
| 32DVSNKGWW | post | 240,2 |
| 33KJJ53U6T | pre | 586,3 |
| 35FFU5UTPJ | pre | 1512,7 |
| 3QSBDDZBUA | pre | 400,0 |
| NSA6OW2VV4 | pre | 1351,0 |
| NSA6OW2VV4 | post | 463,7 |
| PDLAOH6LWY | pre | 645,1 |
| PDLAOH6LWY | post | 395,1 |
| VSUMX3OWK2 | pre | 439,2 |
| VSUMX3OWK2 | post | 493,1 |
| W6DSGFIPDD | pre | 184,3 |
| W6DSGFIPDD | post | 370,6 |
| 2BVT5ZYZGW | pre | 556,9 |
| 2BVT5ZYZGW | post | 532,4 |
| 3ANOEQCO7G | pre | 493,1 |
| 3ANOEQCO7G | post | 571,6 |
| HJQW56FI3S | pre | 1199,0 |
| HJQW56FI3S | post | 444,1 |
| FHL5ZENHKY | pre | 233,3 |
| FHL5ZENHKY | post | 238,2 |
| GDNRS3HZ6W | pre | 189,2 |
| GDNRS3HZ6W | post | 404,9 |
| H2IHUTUID5 | pre | 498,0 |
| H2IHUTUID5 | post | 591,2 |
| KGUDDMHHZZ | pre | 738,2 |
| KGUDDMHHZZ | post | 295,8 |
| MUSL34PBSH | pre | 572,0 |
| MUSL34PBSH | post | 393,4 |
| PXZ7K52TQI | pre | 278,4 |
| PXZ7K52TQI | post | 345,4 |
| YADFULUJ4E | pre | 493,5 |
| YADFULUJ4E | post | 642,9 |

| No rejection, deletion homozygous | Rejection, deletion homozygous | aHUS | No rejection, control sample | Rejection, control sample |
| --- | --- | --- | --- | --- |

aHUS, atypical hemolytic uremic syndrome; CFH, complement factor H; ELISA, Enzyme-linked immunosorbent assay

**Supplementary Table S19. CFH IgG ELISA results for serum samples**

| **Sample ID** | | **Pre/post** | | **CFH IgG (AU/mL)** | | **OD** | |
| --- | --- | --- | --- | --- | --- | --- | --- |
| ABGR4XFUNH | | pre | | 542,2 | | 0,097 | |
| ABGR4XFUNH | | post | | 1100,8 | | 0,168 | |
| BCZORKR4A3 | | pre | | 4862,5 | | 0,650 | |
| DGYMLR5QCF | | pre | | 2553,9 | | 0,354 | |
| EN257NXZIH | | pre | | 1667,2 | | 0,241 | |
| NQZTUVJ6JA | | pre | | 667,2 | | 0,113 | |
| 3RVWTIW47P | | pre | | 2968,0 | | 0,407 | |
| 3RVWTIW47P | | post | | 5538,3 | | 0,736 | |
| ADCKGCMCUK | | pre | | 1210,2 | | 0,182 | |
| ADCKGCMCUK | | post | | 61,7 | | 0,035 | |
| JUIHLJXRHX | | pre | | 690,6 | | 0,116 | |
| JUIHLJXRHX | | post | | 518,8 | | 0,094 | |
| QDM2EBUWCF | | pre | | 1608,6 | | 0,233 | |
| QDM2EBUWCF | | post | | 714,1 | | 0,119 | |
| V53C3LDXJW | | pre | | 2268,8 | | 0,318 | |
| V53C3LDXJW | | post | | 655,5 | | 0,111 | |
| TSMOSHTLU4 | | pre | | 414,4 | | 0,099 | |
| TSMOSHTLU4 | | post | | 889,6 | | 0,195 | |
| ADSAZ3KKXD | | pre | | 577,7 | | 0,132 | |
| 2C26AIVPU6 | | pre | | 862,4 | | 0,189 | |
| 2C26AIVPU6 | | post | | 1656,9 | | 0,350 | |
| 2LQUST2UOF | | pre | | 399,5 | | 0,096 | |
| 32DVSNKGWW | | pre | | 100,0 | | 0,035 | |
| 32DVSNKGWW | | post | | 1,0 | | 0,015 | |
| 33KJJ53U6T | | pre | | 260,9 | | 0,068 | |
| 35FFU5UTPJ | | pre | | 775,7 | | 0,172 | |
| 3QSBDDZBUA | | pre | | 1892,1 | | 0,397 | |
| NSA6OW2VV4 | | pre | | 241,1 | | 0,064 | |
| NSA6OW2VV4 | | post | | 550,5 | | 0,126 | |
| PDLAOH6LWY | | pre | | 340,1 | | 0,084 | |
| PDLAOH6LWY | | post | | 107,4 | | 0,037 | |
| VSUMX3OWK2 | | pre | | 253,5 | | 0,066 | |
| VSUMX3OWK2 | | post | | 185,6 | | 0,051 | |
| W6DSGFIPDD | | pre | | 780,0 | | 0,158 | |
| W6DSGFIPDD | | post | | 513,3 | | 0,110 | |
| AQAS3J2O6O | | pre | | 680,7 | | 0,225 | |
| AQAS3J2O6O | | post | | 890,9 | | 0,281 | |
| S5HBFTQPGV | | pre | | 1379,5 | | 0,410 | |
| S5HBFTQPGV | | post | | 1104,9 | | 0,337 | |
| CO5EXTX4ES | | pre | | 709,1 | | 0,233 | |
| CO5EXTX4ES | | post | | 1493,2 | | 0,440 | |
| L4SB3ASCYV | | pre | | 7004,5 | | 1,895 | |
| L4SB3ASCYV | | post | | 3841,7 | | 1,060 | |
| E3ZJ3IEFYD | | pre | | 671,2 | | 0,223 | |
| E3ZJ3IEFYD | | post | | 773,5 | | 0,250 | |
| WSJNXHVLOO | | pre | | 1237,5 | | 0,372 | |
| WSJNXHVLOO | | post | | 1764,0 | | 0,511 | |
| WIA6YJHZOE | | pre | | 837,9 | | 0,267 | |
| WIA6YJHZOE | | post | | 413,6 | | 0,155 | |
| XTVPEJJ5IJ | | pre | | 659,8 | | 0,220 | |
| XTVPEJJ5IJ | | post | | 586,0 | | 0,200 | |
| MOHTMGGICB | | pre | | 381,4 | | 0,146 | |
| MOHTMGGICB | | post | | 345,5 | | 0,137 | |
| TQOZISNFW6 | | pre | | 1815,2 | | 0,525 | |
| TQOZISNFW6 | | post | | 6211,0 | | 1,685 | |
| LL5OGNNCVV | | pre | | 976,1 | | 0,303 | |
| LL5OGNNCVV | | post | | 531,1 | | 0,186 | |
| VSUMX3OWK2 | | pre | | 803,8 | | 0,258 | |
| VSUMX3OWK2 | | post | | 313,3 | | 0,128 | |
| HHOB2EFYFZ | | pre | | 2201,4 | | 0,671 | |
| HHOB2EFYFZ | | post | | 1115,7 | | 0,367 | |
| ZER4MUBQVQ | | pre | | 1165,7 | | 0,381 | |
| ZER4MUBQVQ | | post | | 2433,6 | | 0,736 | |
| DN7WPTGSG3 | | pre | | 1397,9 | | 0,446 | |
| DN7WPTGSG3 | | post | | 1576,4 | | 0,496 | |
| NI46WIZ4HN | | pre | | 1030,0 | | 0,343 | |
| NI46WIZ4HN | | post | | 1060,4 | | 0,351 | |
| FDYV5RCKXN | | pre | | 710,4 | | 0,253 | |
| FDYV5RCKXN | | post | | 776,4 | | 0,272 | |
| W5J2MOBKEY | | pre | | 551,4 | | 0,209 | |
| W5J2MOBKEY | | post | | 947,9 | | 0,320 | |
| ZN4CIZZGSF | | pre | | 867,5 | | 0,297 | |
| ZN4CIZZGSF | | post | | 1038,9 | | 0,345 | |
| SQVJYO3GQ2 | | pre | | 858,6 | | 0,295 | |
| SQVJYO3GQ2 | | post | | 1080,0 | | 0,357 | |
| 2BVT5ZYZGW | | pre | | 1788,3 | | 0,340 | |
| 2BVT5ZYZGW | | post | | 1230,0 | | 0,239 | |
| 3ANOEQCO7G | | pre | | 493,9 | | 0,107 | |
| 3ANOEQCO7G | | post | | 582,8 | | 0,123 | |
| HJQW56FI3S | | pre | | 1732,8 | | 0,330 | |
| HJQW56FI3S | | post | | 1182,8 | | 0,231 | |
| FHL5ZENHKY | | pre | | 821,7 | | 0,166 | |
| FHL5ZENHKY | | post | | 785,6 | | 0,159 | |
| GDNRS3HZ6W | | pre | | 55,0 | | 0,028 | |
| GDNRS3HZ6W | | post | | 207,8 | | 0,055 | |
| H2IHUTUID5 | | pre | | 505,0 | | 0,109 | |
| H2IHUTUID5 | | post | | 621,7 | | 0,130 | |
| KGUDDMHHZZ | | pre | | 474,4 | | 0,103 | |
| KGUDDMHHZZ | | post | | 263,0 | | 0,068 | |
| MUSL34PBSH | | pre | | 665,4 | | 0,167 | |
| MUSL34PBSH | | post | | 588,2 | | 0,148 | |
| PXZ7K52TQI | | pre | | 702,0 | | 0,176 | |
| PXZ7K52TQI | | post | | 488,6 | | 0,124 | |
| YADFULUJ4E | | pre | | 1915,4 | | 0,475 | |
| YADFULUJ4E | | post | | 844,3 | | 0,211 | |
| cut-off (OD)* | 0,453 | |  | |  | |  |
| No rejection, deletion homozygous | Rejection, deletion homozygous | | aHUS | | No rejection, control sample | | Rejection, control sample |

aHUS, atypical hemolytic uremic syndrome; AU, arbitrary unit; CFH, complement factor H; IgG, immunoglobulin G; ELISA, enzyme-linked immunosorbent assay; OD, optical density

*cut off is calculated according to user’s manual using OD mean and OD standard deviation values

**Supplementary Table S20. Cox proportional hazards model on the association of quartiles of mismatch sum of kidney-related variants and acute rejection.**

| **Covariate** | **Hazard ratio (95% CI)** | **P value** |
| --- | --- | --- |
| Quartiles of mismatch sum of kidney-related variants | 1.13 (0.99-1.28) | 0.071 |
| Recipient age | 0.98 (0.97-0.99) | 0.001 |
| Donor age | 1.03 (1.01-1.04) | <0.001 |
| Recipient sex | 0.82 (0.58-1.14) | 0.235 |
| Donor sex | 1.99 (1.49-2.66) | <0.001 |
| Cold ischemia | 1.00 (1.00-1.00) | 0.089 |
| PRAI | 1.00 (0.99-1.01) | 0.608 |
| PRAII | 1.01 (1.00-1.02) | 0.010 |
| HLAI eplet mismatch | 1.01 (0.99-1.03) | 0.460 |
| HLAII eplet mismatch | 1.03 (1.02-1.04) | <0.001 |

CI, confidence interval; HLA, human leukocyte antigen; PRA, panel reactive antibody

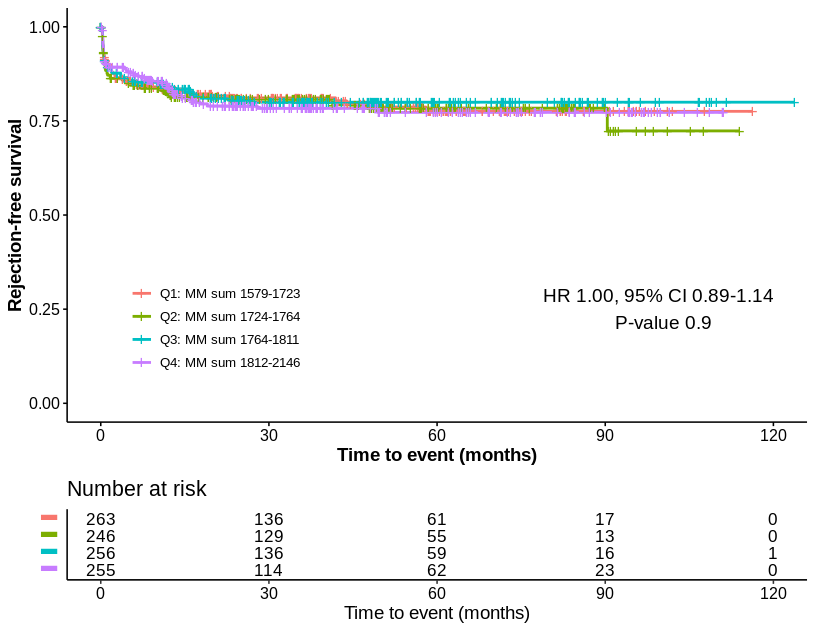

**Supplementary Figure S1. The effect of quartiles of missense variant mismatch sum coding for transmembrane and secretory proteins on rejection-free graft survival.** The quartile 1 (Q1) represents the lowest number of mismatch between recipient and donor, quartile 4 (Q4) the highest number of mismatch. An event (acute rejection) occurs each time the curve drops. The tick marks indicate censored data (end of follow-up time). Unadjusted hazard ratio (HR) with confidence interval (CI) shown. CI, confidence interval; HR, hazard ratio; MM, mismatch

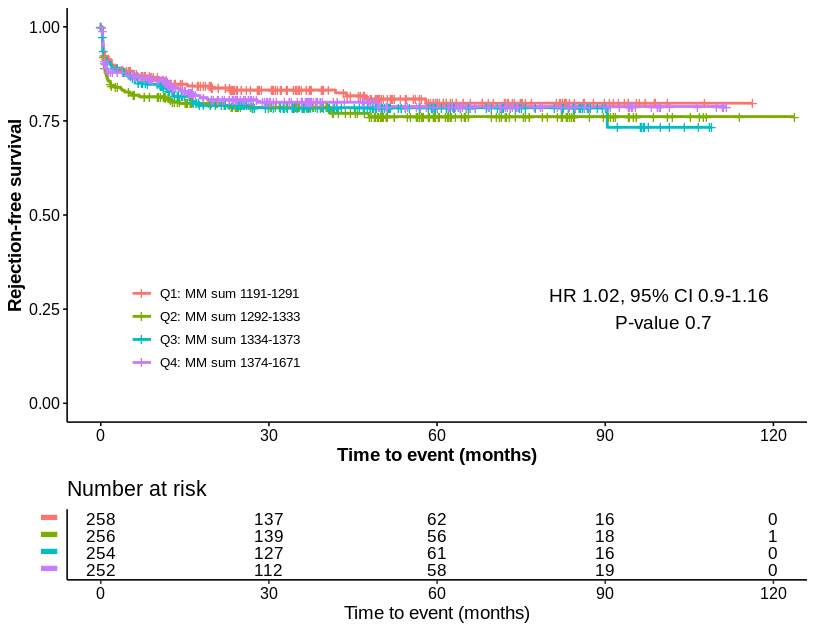
**Supplementary Figure S2. The effect of quartiles of missense variant mismatch sum coding for transmembrane proteins on rejection-free graft survival.** The quartile 1 (Q1) represents the lowest number of mismatch between recipient and donor, quartile 4 (Q4) the highest number of mismatch. An event (acute rejection) occurs each time the curve drops. The tick marks indicate censored data (end of follow-up time). Unadjusted hazard ratio (HR) with confidence interval (CI) shown. CI, confidence interval; HR, hazard ratio; MM, mismatch

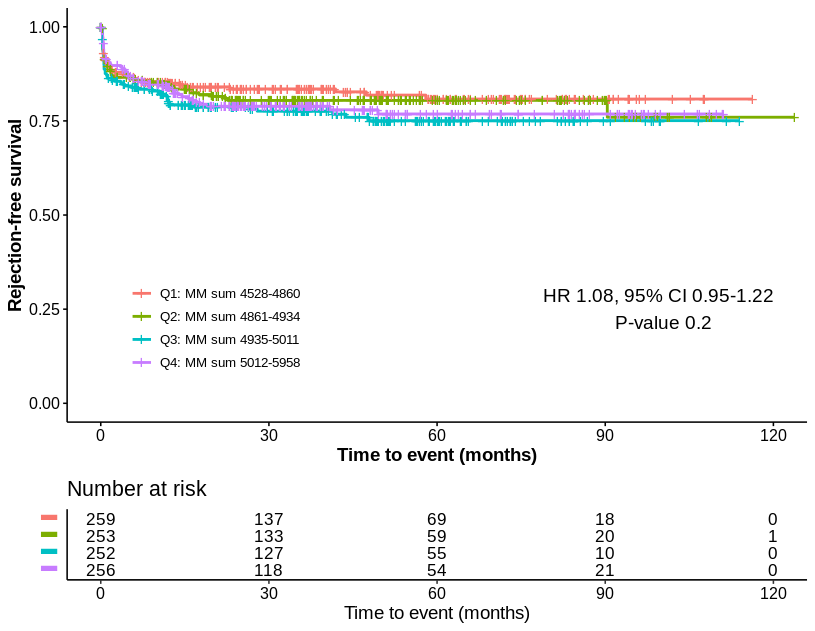

**Supplementary Figure S3. The effect of quartiles of the overall genome-wide missense variant mismatch sum on rejection-free graft survival.** The quartile 1 (Q1) represents the lowest number of mismatch between recipient and donor, quartile 4 (Q4) the highest number of mismatch. An event (acute rejection) occurs each time the curve drops. The tick marks indicate censored data (end of follow-up time). Unadjusted hazard ratio (HR) with confidence interval (CI) shown. CI, confidence interval; HR, hazard ratio; MM, mismatch

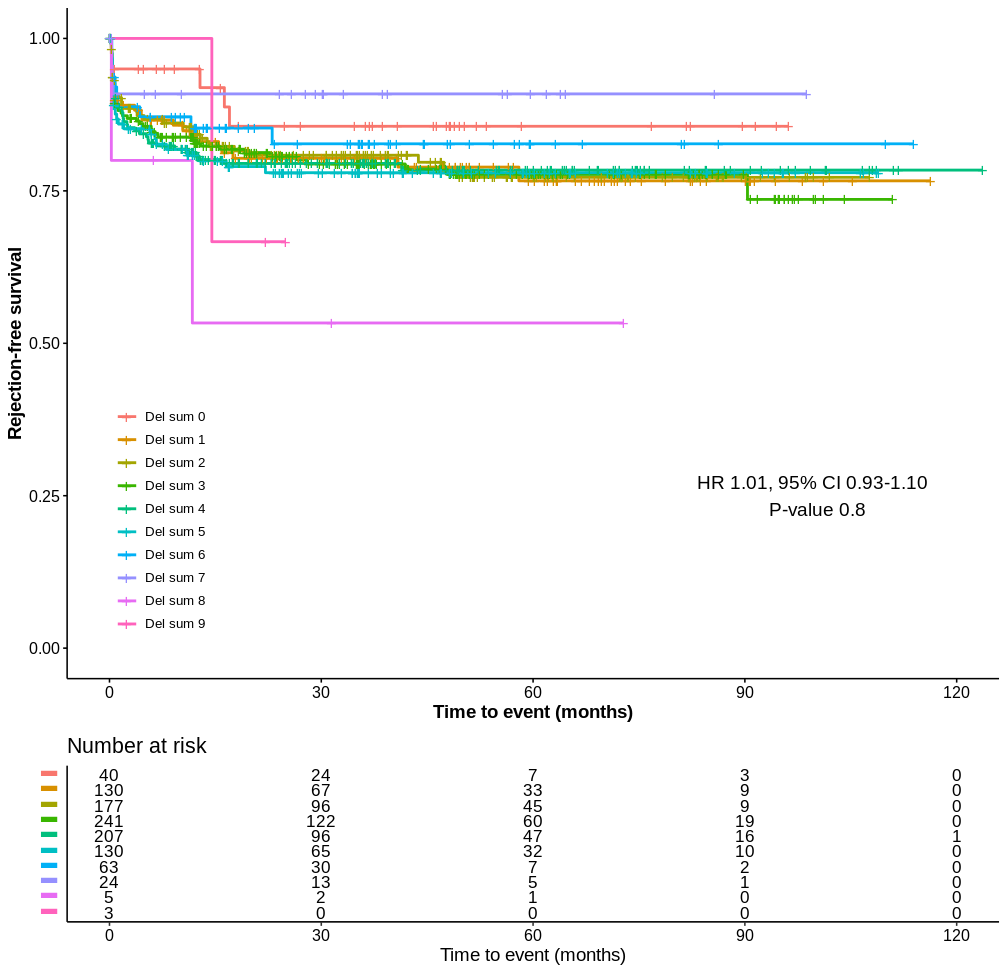

**Supplementary Figure S4. The effect of deletion mismatch sum on rejection-free graft survival in donor-recipient pairs.** In the donor-recipient analysis of mismatch sum, the recipient who was homozygous for a deletion-tagging allele received a transplant from a donor with a non-homozygous genotype. The sum of mismatches of deletion-tagging alleles was calculated and ranged between 0-9. An event (acute rejection) occurs each time the curve drops. The tick marks indicate censored data (end of follow-up time). Unadjusted hazard ratio (HR) with confidence interval (CI) shown. CI, confidence interval; Del, deletion; HR, hazard ratio

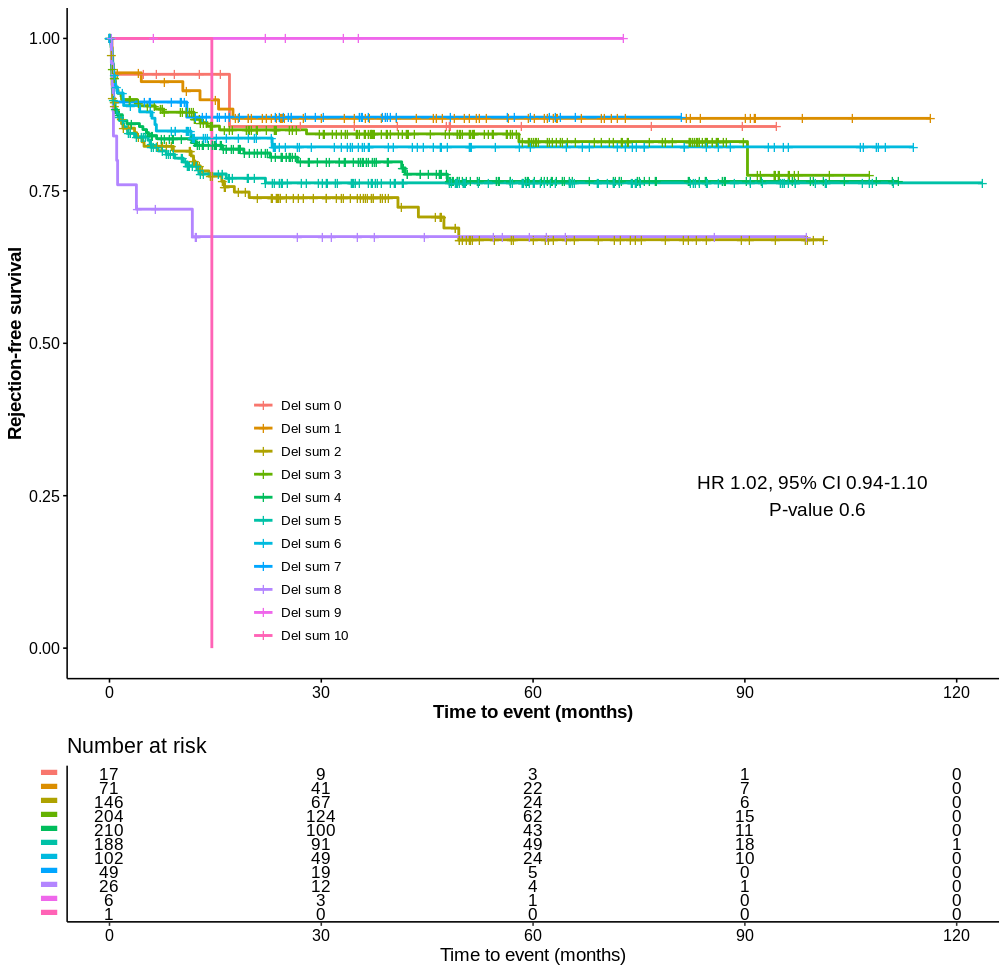

**Supplementary Figure S5. The effect of deletion sum on rejection-free graft survival among recipients.** In recipient-only analysis of deletion sum, the sum of deletions was calculated on the basis of homozygous genotype of deletion-tagging allele. The sum of deletions among recipients ranged between 0-10. An event (acute rejection) occurs each time the curve drops. The tick marks indicate censored data (end of follow-up time). Unadjusted hazard ratio (HR) with confidence interval (CI) shown. CI, confidence interval; Del, deletion; HR, hazard ratio

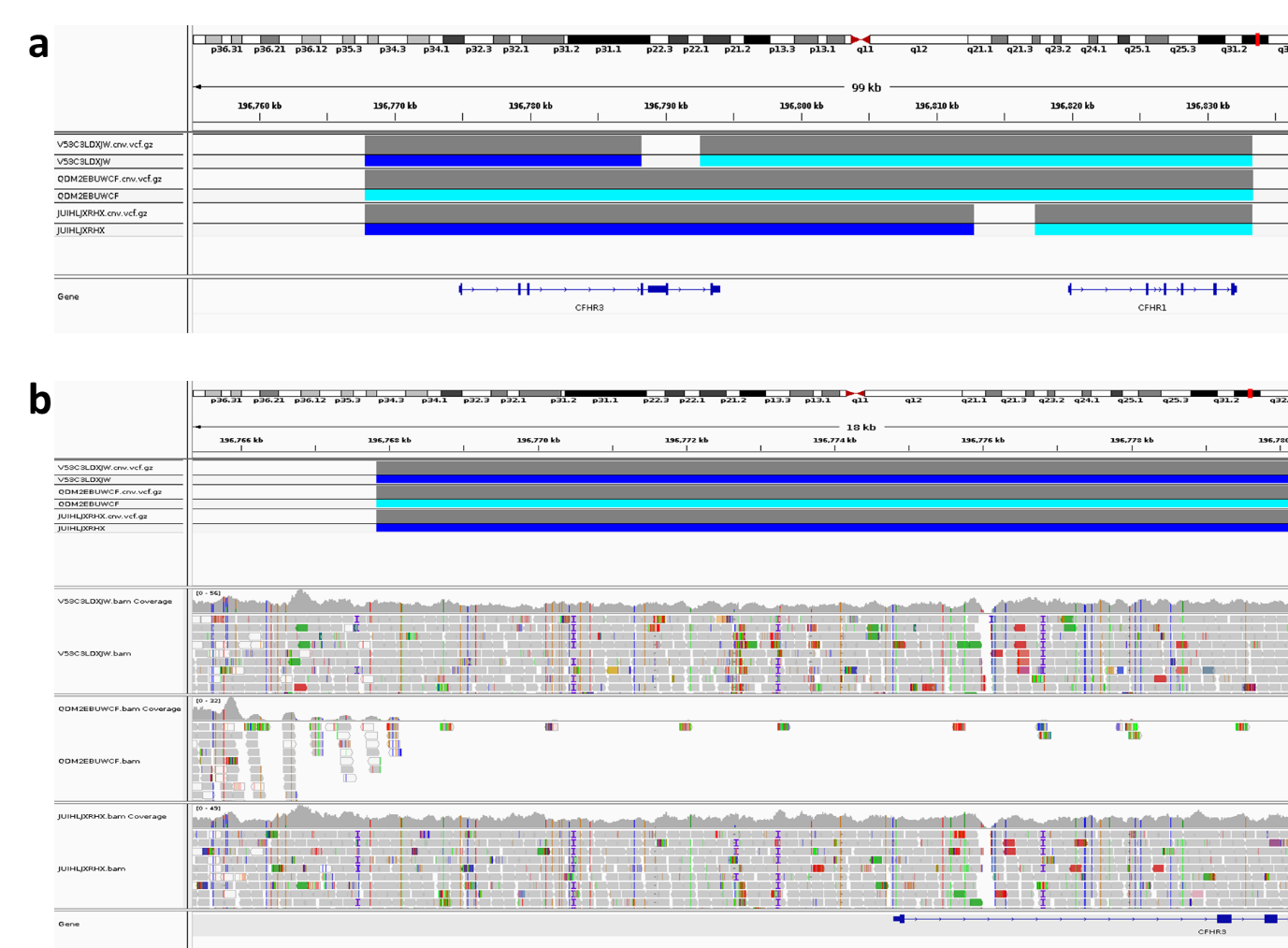

**Supplementary Figure S6. The results of whole genome sequencing of CFH- and CFH-related protein loci on chromosome 1.** The upper panel A shows the length of deletions in CFHR3-CFHR1 region on chromosome 1 for each three samples. The light blue color represents a homozygous deletion, and the dark blue heterozygous deletion. The lower panel B shows the lack of reads in the homozygous region of one sample in the middle.
